# Routes of importation and spatial dynamics of SARS-CoV-2 variants during localised interventions in Chile

**DOI:** 10.1101/2024.01.18.24301504

**Authors:** Bernardo Gutierrez, Joseph L.-H. Tsui, Giulia Pullano, Mattia Mazzoli, Karthik Gangavarapu, Rhys P.D. Inward, Sumali Bajaj, Rosario Evans Pena, Simon Busch-Moreno, Marc A. Suchard, Oliver G. Pybus, Alejandra Dunner, Rodrigo Puentes, Salvador Ayala, Jorge Fernandez, Rafael Araos, Leo Ferres, Vittoria Colizza, Moritz U.G. Kraemer

**Affiliations:** Department of Biology, University of Oxford, Oxford, UK; Colegio de Ciencias Biológicas y Ambientales, Universidad San Francisco de Quito USFQ, Quito, Ecuador; Department of Biology, Georgetown University, Washington D.C., USA; INSERM, Sorbonne Université, Institut Pierre Louis d’Epidémiologie et de Santé Publique, IPLESP, Paris, France; ISI Foundation, Turin, Italy; Department of Human Genetics, University of California Los Angeles, Los Angeles, CA, USA; Department of Biostatistics, University of California Los Angeles, Los Angeles, CA, USA; Department of Biomathematics, University of California Los Angeles, Los Angeles, CA, USA; Pandemic Sciences Institute, University of Oxford, UK; Department of Pathobiology and Population Science, Royal Veterinary College, London, UK; Instituto de Salud Pública de Chile, Santiago, Chile; Instituto de Ciencias e Innovación en Medicina (ICIM), Facultad de Medicina Clínica Alemana, Universidad del Desarrollo, Santiago, Chile; Data Science Institute, Universidad del Desarrollo, Santiago, Chile; Telefónica, Santiago, Chile; Tokyo Tech World Research Hub Initiative, Institute of Innovative Research, Tokyo Institute of Technology, Tokyo, Japan.

**Keywords:** Genomic epidemiology, viral importations, spatial invasion

## Abstract

South America suffered large SARS-CoV-2 epidemics between 2020 and 2022 caused by multiple variants of interest and concern, some causing substantial morbidity and mortality. However, their transmission dynamics are poorly characterised. The epidemic situation in Chile enables us to investigate differences in the distribution and spread of variants Alpha, Gamma, Lambda, Mu and Delta. Chile implemented non-pharmaceutical interventions and an integrated genomic and epidemiological surveillance system that included airport and community surveillance to track SARS-CoV-2 variants. Here we combine viral genomic data and anonymised human mobility data from mobile phones to characterise the routes of importation of different variants into Chile, the relative contributions of airport-based importations to viral diversity versus land border crossings and test the impact of the mobility network on the diffusion of viral lineages within the country. We find that Alpha, Lambda and Mu were identified in Chile via airport surveillance six, four and five weeks ahead of their detection via community surveillance, respectively. Further, some variants that originated in South America were imported into Chile via land rather than international air travel, most notably Gamma. Different variants exhibited similar trends of viral dissemination throughout the country following their importation, and we show that the mobility network predicts the time of arrival of imported lineages to different Chilean comunas. Higher stringency of local NPIs was also associated with fewer domestic viral importations. Our results show how genomic surveillance combined with high resolution mobility data can help predict the multi-scale geographic expansion of emerging infectious diseases.

**Significance statement:** Global preparedness for pandemic threats requires an understanding of the global variations of spatiotemporal transmission dynamics. Regional differences are important because the local context sets the conditions for the unfolding of local epidemics, which in turn affect transmission dynamics at a broader scale. Knowledge gaps from the SARS-CoV-2 pandemic remain for regions like South America, where distinct sets of viral variants emerged and spread from late 2020 onwards, and where changes in human behaviour resulted in epidemics which differed from those observed in other regions. Our interdisciplinary analysis of the SARS-CoV-2 epidemic in Chile provides insights into the spatiotemporal trends of viral diffusion in the region which shed light on the drivers that can influence future epidemic waves and pandemics.

## Main text

The epidemic dynamics of SARS-CoV-2 are context-dependent. Intensity of transmission has been shown to vary greatly due to a combination of factors related to population mixing, international travel, socio-economic indicators, and SARS-CoV-2 lineage composition (1–3). The emergence of novel variants of interest (VOIs) and variants of concern (VOCs) were typically associated with epidemic waves in the country of their first report and later caused epidemics either regionally (Gamma and Beta in Brazil and South Africa, respectively) or globally (Alpha, Delta and Omicron in England, India and South Africa, respectively; (1, 4–7)).

During the emergence of Gamma, Beta, and Alpha in late 2020, the likelihood of a variant being detected in a country could be predicted by the number of passengers arriving from the countries reporting initial outbreaks (8). During that time, international and local travel was still restricted compared to pre-pandemic levels (9). Studies have shown how increased rates of local mixing impacted SARS-CoV-2 growth rates and total numbers of cases (10, 11) and suggest that different types of human mobility contributed to epidemic spread (12, 13).

Despite these general insights, the transmission dynamics of multiple variants in Latin America remain understudied and it is still unknown why some variants spread in this region and others did not. Multiple factors likely contributed to SARS-CoV-2 lineage composition (14), including high transmission rates during the times when some variants emerged: the Gamma variant in the Brazilian Amazon (5), the Lambda variant in Peru and Chile (15), and the Mu variant in Colombia (16, 17). Various other lineages not designated as VOIs or VOCs in regions like Mexico (18) and the USA (e.g. Iota and Epsilon; (19–21) were also detected in the continent but did not reach high frequencies.

Here we analyse 787 SARS-CoV-2 genomes collected via dedicated airport genomic surveillance at Santiago de Chile Airport (SCL) and 7957 genomes generated through community genomic surveillance (22, 23). This data is combined with human mobility data from mobile phones (covering 24.3% (24) of Chile’s mobile phone subscribers) and records of international arrivals into the country (both airport arrivals and total land border crossings during 2021) to explore how international arrivals, airport testing, and local interventions (implemented at the lowest administrative divisions in the country, *comunas*) impacted the transmission dynamics of SARS-CoV-2 lineages in Chile during 2020-2021. We also assess how Chile’s mobility network predicts viral spread following new importations. While similar analyses have been performed for other locations and contexts, Chile provides a distinct scenario to study these phenomena. Its genomic surveillance programme at the largest international airport in the country (that received the vast majority of international travellers which were all tested upon arrival during 2021), combined with its network of land international ports of entry across its large border with Argentina (and smaller borders with Bolivia and Peru), result in an ideal scenario to evaluate the routes by which individual variants were imported into the country. Also, the highly localised application of non-pharmaceutical interventions (NPIs) during the pandemic (25–28) were a unique phenomenon which has not been evaluated in its efficacy in limiting the geographic spread of SARS-CoV-2.

### The SARS-CoV-2 landscape in Chile and South America in 2021

Different SARS-CoV-2 variants emerged in South America during late 2020 and early 2021 (29). The earliest VOC described in South America was the Gamma variant, which emerged in Manaus, Brazil (5) and quickly spread to the southern half of the continent, into Argentina, Chile, Uruguay, Bolivia and Paraguay (**Fig. 1A**). Simultaneously the Lambda variant emerged and rapidly increased in frequency in Peru and Chile, while the Mu variant appeared later, around April 2021, in Colombia, Ecuador and Venezuela (**Fig. 1A**). The epidemiological trends across different countries during this time suggest that these emerging variants drove national epidemic peaks, as increases in the number of cases followed increases in the prevalence of each variant between February and May 2021 across the region. Further epidemic peaks followed in June and July, which might have been driven by viral lineage replacement caused by regional movements of various South American variants or by the importation of the Delta variant, which swept across the world from spring 2021 onwards (**Fig. 1A**). Variants that became dominant in some other regions of the world, like Alpha and Beta, did not dominate in South America (**Fig. S1**).

**Figure 1.**
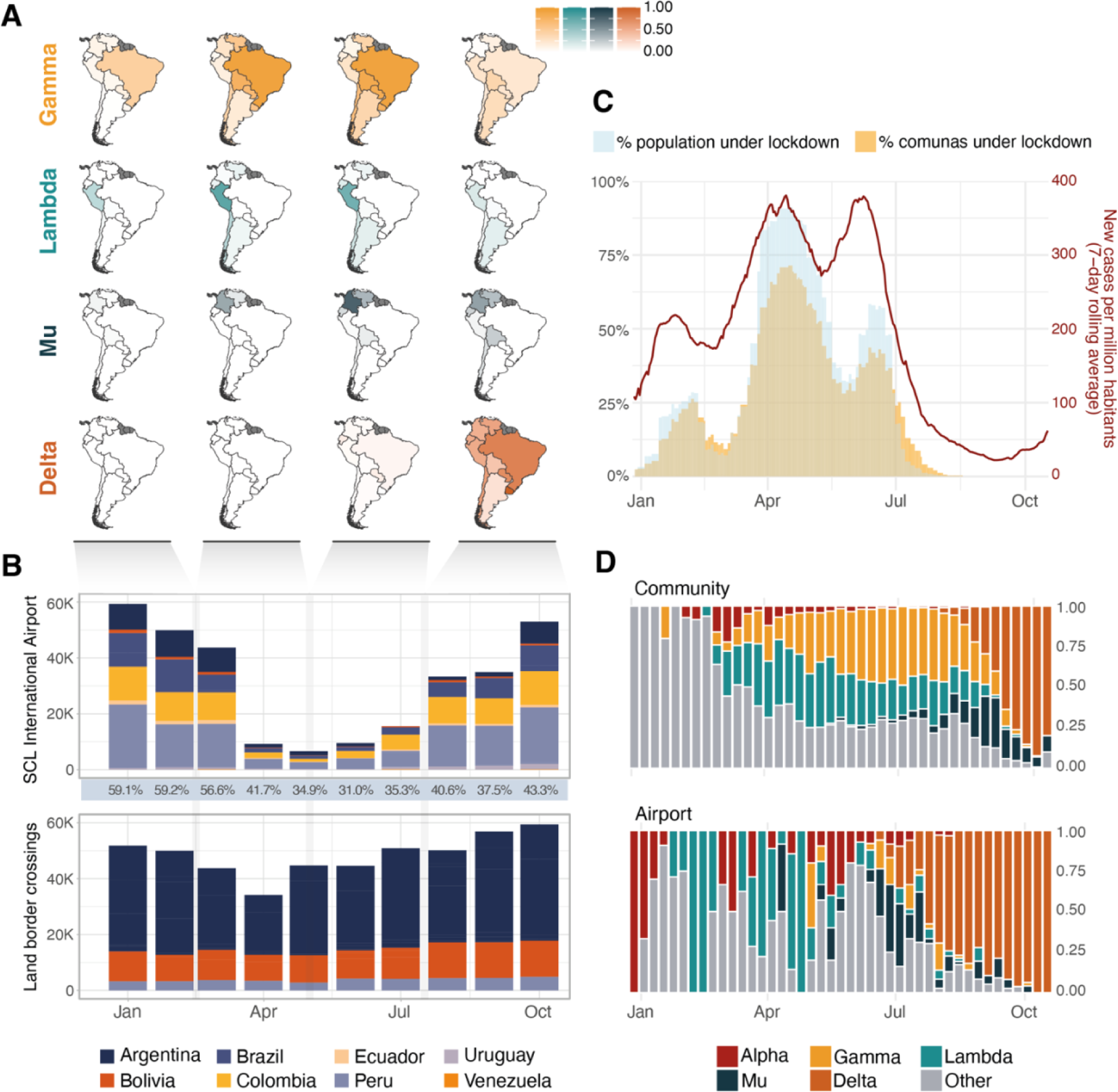
SARS-CoV-2 in Chile and South America during 2021. **A)** Relative frequency of four significant VOIs/VOCs in South American countries, shown as the number of sequences from each variant relative to the total genomic surveillance output from each country over the specified time period. **B)** Total volume of passengers from South American countries arriving in Chile via the Santiago de Chile International Airport (SCL) and through land border crossings from neighbouring countries during 2021. Percentages under the upper panel show the proportion of all arrivals that come from South American countries vs the rest of the world. **C)** Epidemiological COVID-19 trends during 2021 showing the total number of cases reported in the country, and the number of comunas and people in the country placed under lockdown over time; the latter consider comunas under the highest lockdown tier, full lockdown, which restricted mobility every day of the week within the comuna. **D)** Proportions of VOIs and VOCs detected in Chile during 2021 under their two surveillance schemes, community surveillance (top) and airport surveillance of international travellers arriving at SCL (bottom).

These variant dynamics across other countries and their connectivity to Chile likely played a role on the country’s SARS-CoV-2 epidemic trends (**Fig. 1B-D**). Data on monthly land border crossings and international airport arrivals shows that both Peru and Argentina were important contributors to the total incoming travellers to Chile during 2021, albeit with different trends; while Peru accounted for the majority of air travellers arriving every month via SCL (up to 22.7% of all air passengers per month, **Fig. 1B**), more total travellers arrived from Argentina through land travel (14 land border crossings with Chile were operationally active during 2021, which recorded between 67% and 100% of all incoming travellers from Argentina into the country each month; **Fig. 1B**). We note that travellers from Argentina increased considerably from May 2021 onwards possibly reducing the impact of air travel restrictions in the flow of people from that country during that time (specifically May, June and July 2021; **Fig. 1B**).

During the study period between November 2020 and October 2021, Chile experienced three epidemic waves, peaking in February, April and July, respectively. Each wave was followed by periods during which individual comunas enacted lockdowns of different stringency (i.e. the degree to which people could move freely; **Supporting Text**) and were implemented as a response to the changing epidemic trends. This resulted in a fluctuation of the number of comunas and total people under restrictions over time (**Fig. 1C**).

### International importations of viral lineages depend on country-level prevalence and human mobility

Chile implemented separate genomic surveillance programmes for incoming international travellers at SCL and within the community. The proportions of different variants in both datasets show that for Alpha, Lambda, Mu and Delta, airport detections preceded the detection of these variants in the community by six, four, five and two weeks respectively. Gamma however was rarely observed using data from airport surveillance yet consistently exceeded 25% of sequences from cases sampled in the community from 25 April 2021 until the end of August, when Delta started to replace all other lineages (**Fig. 1D**).

From these observations, we hypothesise that variants like Gamma were imported to Chile through different pathways other than international air travel. We performed phylogenetic analysis of virus genomic data from all five variants to map the number and timing of importations in Chile between late 2020 and throughout 2021 and found frequent and sustained importations across the study period. Alpha and Lambda importations peaked simultaneously around April while Gamma saw sustained importations between May and August 2021. Mu had a peak in importations around July, followed by large numbers of Delta importations between August and October (**Fig. 2A**). Phylodynamic analysis of each variant reveals an estimated ∼648 independent introductions (excluding introductions of all other non-VOI/VOC viral lineages). For Delta we detect the highest number of importations (median 336 importations; 95% HPD: 327-344), considerably higher than for Lambda (median 111 importations; 95% HPD: 95-124), Mu (median 86 importations; 95% HPD: 79-92), Gamma (median 74 importations; 95% HPD: 64-83) and Alpha (median 41 importations; 95% HPD: 35-43; **Fig. 2B**). These estimates are likely affected by sampling intensity heterogeneities across countries and by the limited proportion of sampled cases in Chile, which also exhibit local sampling variation across space and time. We note a general trend whereby an increasing proportion of cases were sequenced coincided with declining numbers of cases (**Fig. S2**). We accounted for these potential biases by using a mobility-informed subsampling approach that considers human movements from international destinations into Chile (see *Methods* and (18)).

**Figure 2.**
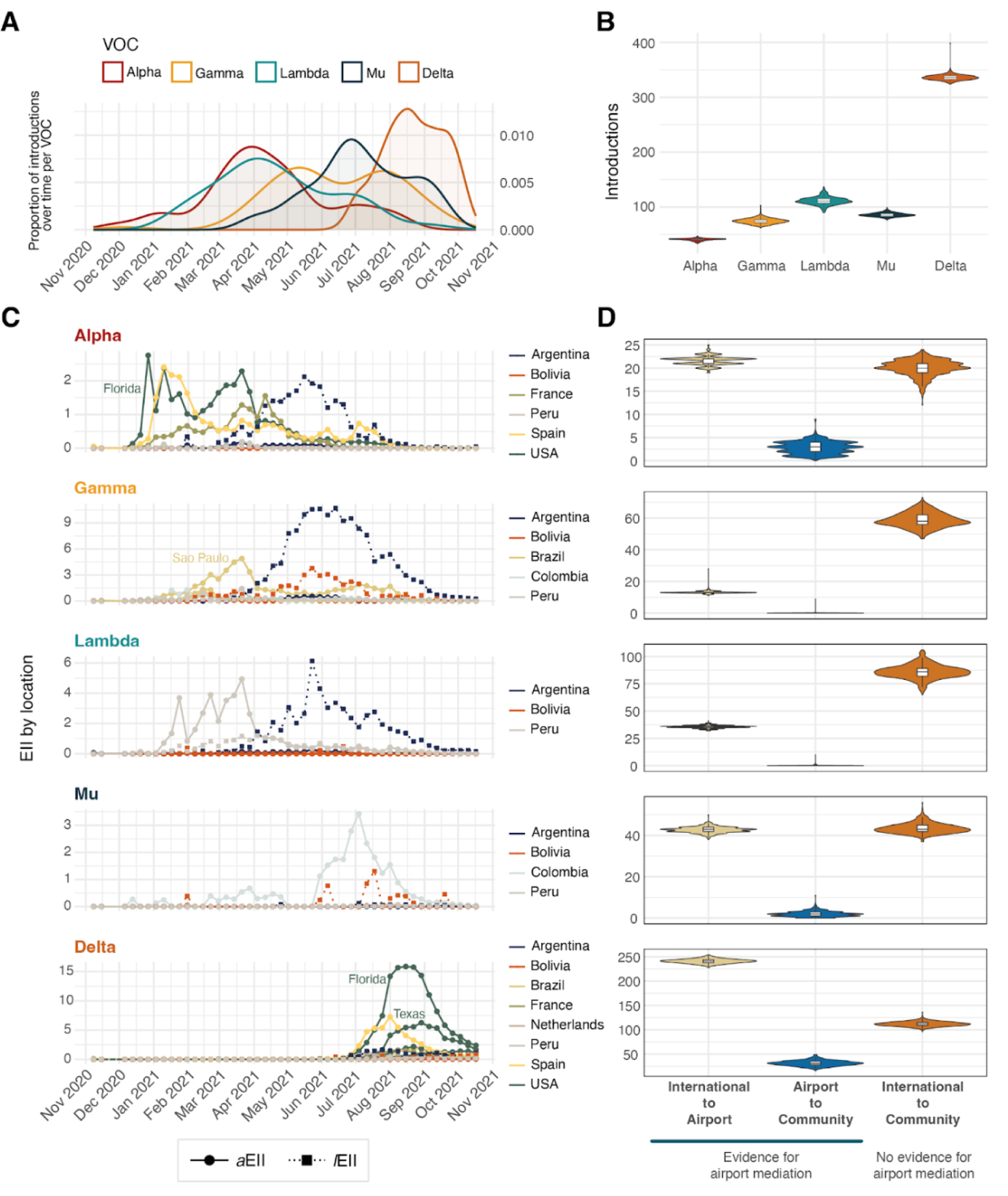
Viral importation dynamics and ports of entry. **A)** Trends in inferred viral importations over time. Smoothed density estimates of the numbers of importations per variant over time plotted by the TMRCA of individual transmission lineages. **B)** Posterior probability densities of the inferred number of viral importations per variant estimated through Bayesian phylogeographic analysis. **C)** Time series for the estimated importation intensity (EII) indices from selected states (for USA and Brazil - described in each panel) and countries. EIIs are estimated weekly for each variant and country; solid lines with circles show EIIs based on air travel volume (aEII) and dashed lines with squares show EIIs based on land border crossings (lEII). D) Posterior probability densities of the inferred number of importations stratified by importation route per variant. ‘International to Airport’ show estimated transitions from international location nodes to SCL airport nodes; ‘Airport to Community’ show estimated transitions from international location nodes to SCL airport nodes to Chile nodes detected through community surveillance; ‘International to Community’ show estimated transitions from international location nodes to Chile nodes detected through community surveillance.

An estimated 43% of introductions led to detectable onward transmission in Chile, hereafter called “transmission lineages” (TLs). As with previous studies, the size distribution of these TLs is heavily skewed, with a small portion of importations leading to large TLs while the vast majority resulted in limited detectable onward transmission (**Fig. S3**); this size distribution might be affected by the study period cutoff date, as the persistence of TLs over time is right-censored (**Fig. S4**).

Because phylogenetic inferences are influenced by sampling biases and model specification we use independent datasets to correlate estimates made from genomic data. Previous studies have shown that the number of international travellers coming into a territory can be used to help infer the expected number of SARS-CoV-2 case introductions (30, 31). We calculated an estimated importation intensity index (EII) based on the estimated cases for each variant in a potential source country and the total movements from that country into Chile. We used a Granger-causality (GC) test to ask whether EIIs can ‘forecast’ the inferred importations of each variant over time (see *Methods*). We found that the weekly number of viral importations inferred from genomic data followed the EII for each variant (Table 1). Interestingly, Gamma introductions are best predicted by an EII that only includes land-based border crossings (*l*EII) while the remaining four variants are predicted by an EII based on air-based human movements (*a*EII) rather than land border crossings (**Table 1**). These results may explain why Gamma was rarely detected during airport genomic surveillance.

**Table 1.** Granger-causality test results between phylogenetically inferred viral imports and estimated importation intensity indices for both air (aEII) and land (lEII) mobility components.

| VOC | Source location | $\alpha$ EII | | | /EII | | |
| --- | --- | --- | --- | --- | --- | --- | --- |
|  |  | Lag (model support) | F statistic | p value | Lag (model support) | F statistic | p value |
| *Alpha* | Florida (USA) | 12 (AIC = -6.74) | 10.65 | 0.001 | --- | --- | --- |
|  | France | 3 (SC = -2.65) | 7.47 | 0.0005 | --- | --- | --- |
|  | Spain | 12 (AIC = -6.83) | 1.77 | 0.21 | --- | --- | --- |
|  | *Argentina* | 12 (AIC = -9.89) | 7.42 | 0.001 | 1 (SC = -2.24) | 0.04 | 0.84 |
|  | *Bolivia\*\** | --- | --- | --- | --- | --- | --- |
|  | *Peru* | 2 (SC = -5.84) | 8.95 | 0.000594 | 1 (SC = -8.53) | 14.83 | 0.0004 |
|  | **Combined\*** | 12 (AIC = 3.42) | 20.98 | 0.0001 | 1 (SC = -2.24) | 0.07 | 0.79 |
| *Gamma* | Rio de Janeiro (BRA) | 6 (SC = -4.29) | 0.39 | 0.88 | --- | --- | --- |
|  | Santa Catarina (BRA) | 11 (SC = -14.99) | 1.07 | 0.46 | --- | --- | --- |
|  | Sao Paulo (BRA) | 1 (SC = 0.47) | 0.03 | 0.86 | --- | --- | --- |
|  | Colombia | 12 (SC = -3.31) | 1.00 | 0.52 | --- | --- | --- |
|  | *Argentina* | 1 (SC = -3.62) | 3.50 | 0.07 | 1 (SC = 1.63) | 11.14 | 0.002 |
|  | *Bolivia* | 1 (SC = -6.05) | 0.44 | 0.51 | 1 (SC = 0.52) | 3.61 | 0.06 |
|  | *Peru* | 1 (SC = -1.19) | 0.17 | 0.68 | 1 (SC = -3.91) | 5.75 | 0.02 |
|  | **Combined\*** | 1 (SC = 8.36) | 0.03 | 0.86 | 1 (SC = 2.29) | 9.80 | 0.003 |
| *Lambda* | *Argentina* | 1 (SC = -4.36) | 1.98 | 0.17 | 1 (SC = 1.39) | 0.009 | 0.92 |
|  | *Bolivia* | 1 (SC = -11.22) | 0.53 | 0.47 | 1 (SC = -3.02) | 0.04 | 0.85 |
|  | *Peru* | 10 (SC = -1.95) | 3.84 | 0.01 | 1 (SC = -2.84) | 53.21 | 4.24E-09 |
|  | **Combined\*** | 10 (SC = 5.21) | 3.68 | 0.01 | 1 (SC = 1.42) | 0.64 | 0.43 |
| *Mu* | Colombia | 1 (SC = 0.52) | 7.04 | 0.01 | --- | --- | --- |
|  | *Argentina* | 1 (SC = -1.11) | 0.17 | 0.68 | 1 (SC = -4.77) | 1.19 | 0.28 |
|  | *Bolivia* | 2 (SC = -7.25) | 1.00 | 0.38 | 2 (SC = -0.64) | 1.57 | 0.22 |
|  | *Peru* | 12 (SC = -7.80) | 5.08 | 0.006 | 10 (SC = -9.39) | 3.27 | 0.02 |
|  | **Combined\*** | 1 (SC = 7.54) | 7.50 | 0.009 | 2 (SC = -0.61) | 1.82 | 0.18 |
| *Delta* | Sao Paulo (BRA) | 12 (SC = -16.32) | 1459.4 | 5.47E-12 | --- | --- | --- |
|  | Florida (USA) | 12 (SC = -8.71) | 321.79 | 2.28E-09 | --- | --- | --- |
|  | Georgia (USA) | 12 (SC = -15.77) | 873.46 | 4.25E-11 | --- | --- | --- |
|  | New York (USA) | 12 (SC = -54.78) | 9.93E+15 | 2.20E-16 | --- | --- | --- |
|  | Texas (USA) | 12 (SC = -16.01) | 3553.6 | 1.56E-13 | --- | --- | --- |
|  | France | 12 (SC = -89.96) | 122.5 | 1.06E-07 | --- | --- | --- |
|  | Netherlands | 12 (SC = -8.89) | 449.03 | 6.05E-10 | --- | --- | --- |
|  | Spain | 12 (SC = -7.68) | 188.35 | 1.92E-08 | --- | --- | --- |
|  | *Argentina* | 12 (SC = -6.04) | 2.32 | 1.20E-01 | 12 (SC = -3.71) | 102.46 | 1.71E-09 |
|  | *Bolivia* | 9 (SC = -6.71) | 3.37E+26 | 2.20E-16 | 9 (SC = -64.23) | 3.44E+27 | 2.20E-16 |
|  | *Peru* | 12 (SC = -71.38) | NA*** | NA*** | 12 (SC = -73.92) | NA*** | NA*** |
|  | **Combined\*** | 12 (SC = 4.35) | 8.43 | 0.003 | 12 (SC = -4.05) | 121.25 | 6.87E-10 |

Countries with italicized names share a land border with Chile. Non-neighbouring countries included based on at least one epidemiological week where EII > 1. Cells in orange show significant results at $\alpha = 0.01$ .
\* Refers to $\alpha$ EII estimated from all countries with direct flights into Chile (including those minor contributors not shown in the table) and / EII estimates from all neighbouring countries.
\*\* No genomic surveillance data for this line.
\*\*\* Unavailable test result due to multicollinearity

Airport testing and genomic surveillance were implemented as a public health measure and were combined with requirements for returning travellers of self-isolation following a positive test result (**Supporting Text**). Therefore, this surveillance scheme aimed to minimise and ideally contain transmission from infected incoming travellers. Its effectiveness depends on the sensitivity and accuracy of the testing itself and the proportion of viral introductions through the airport rather than by other routes. We consequently hypothesise that cases identified during airport surveillance would result in more limited transmission in the community. To explore this hypothesis, we used a discrete phylogeographic approach to estimate the number of viral movements between countries other than Chile and the SCL International Airport, the movements from SCL airport into the community within Chile, and importations from other countries directly into community circulation in Chile (i.e., not detected or mediated via airport surveillance). The importation dynamics appear specific for individual variants: Gamma and Lambda importations were predominantly inferred directly from international destinations into the community with no evidence of airport mediation, while Alpha and Mu show an equivalent number of importations into the SCL International Airport as they did into the community. Delta shows a higher proportion of importations through the airport compared to directly into the community. Overall, negligible numbers of transitions from the airport into the community were inferred (**Fig. 2D**). We note however that importations with no evidence of airport mediation could still have been imported through the airport but went undetected due to the timing of testing (no detectable virus at arrival) or that positive cases were not sequenced (although ∼95% of positive cases from airport surveillance during 2021 were sequenced, those excluded were those where sample quality prohibited genome sequencing and likely occurred at random across the collection of samples).

An independent estimation of both *l*EII and *a*EII for individual source countries per variant shows the likely contributors of viral importations over time (**Fig. 2C**), with Gamma and Mu introductions likely derived from a single source country (Argentina and Colombia, respectively), while Alpha and Delta likely being imported from multiple countries (or states within the USA and Brazil) simultaneously. Lambda exhibits a distinct pattern, in that its predominant source changed from Peru (via air travel) during early 2021, to Argentina (via land border crossings) after April 2021 (**Fig. 2C**), although the expected importation wave from Argentina was not observed using genomic data (**Fig. 2A**, **Table 1**). In the case of Gamma, only Argentina and Peru *l*EIIs show a significant correlation with inferred viral introductions, further suggesting that land mobility played a bigger role in the seeding of this variant in Chile compared to air travel. This is corroborated by the increased number of Gamma viral imports directly into the community compared to the airport-mediated importations (**Fig. 2D**). Interestingly, both the *a*EII and *l*EII for Peru significantly correlate with the observed importations for all variants, suggesting that both land and air routes of entry played an important role in how seeding events from this country into Chile took place (despite the fact that there is a single land border crossing point between Peru and Chile; **Table 1**). Again, the relatively high number of introductions that show no evidence of being airport-mediated (**Fig. 2D**) for Alpha, Lambda and Mu suggest that land importations were also commonplace during this time. All global EII estimates (combining both *l*EII and *a*EII, **Fig. S5**) and the phylogenetically inferred viral imports are temporally associated (FAlpha = 15.00, *p*Alpha < 0.001; FGamma = 7.19, *p*Gamma = 0.01; FLambda = 12.97, *p*Lambda < 0.001; FMu = 5.46, *p*Mu = 0.02; FDelta = 32.28, *p*Delta < 0.001).

### Human mobility drives SARS-CoV-2 spatial invasion across comunas

Given the limited ports of entry for different variants into Chile, the arrival of individual lineages to new comunas after their introduction is expected to be driven by infected people moving within the country and seeding new local epidemics. We estimate the impact of local scale human mobility on the invasion dynamics of different TLs by comparing comuna-to-comuna arrival times extracted from a continuous phylogeographic approach (32) (see *Methods*). The spread of these TLs took place during periods of changing mobility in the country as lockdowns were enacted as part of the *Paso a paso* plan, which established a tiered system for the implementation of mobility restrictions to mitigate viral transmission (the plan employed various stringency tiers that can be summarised as (i) a high stringency full lockdown with mobility restrictions every day of the week, a (ii) mid-stringency weekend lockdown with mobility restrictions only during weekends and (iii) a low stringency lockdown with no mobility restrictions; **Supporting Text**). This plan gave individual comunas the authority to determine the appropriate stringency for these NPIs and the appropriate times to change stringency. Consequently, real-time mobility data (estimated from mobile phone data) is important in accurately estimating the contribution of human mobility to viral spread. Between January and October, there were at least three periods when increasing numbers of comunas were placed under lockdown (**Fig. 1C**); an accompanying reduction in mobility followed, reaching its lowest point around the April lockdown season, when up to 80% of comunas were under lockdown (**Fig. 3A**). The bounceback following this downward trend also coincided with a policy implemented on May 26, when individuals with a complete immunisation schedule were issued a “mobility pass” which allowed them free movement, even within comunas under lockdown (**Fig. 3A**).

**Figure 3.**
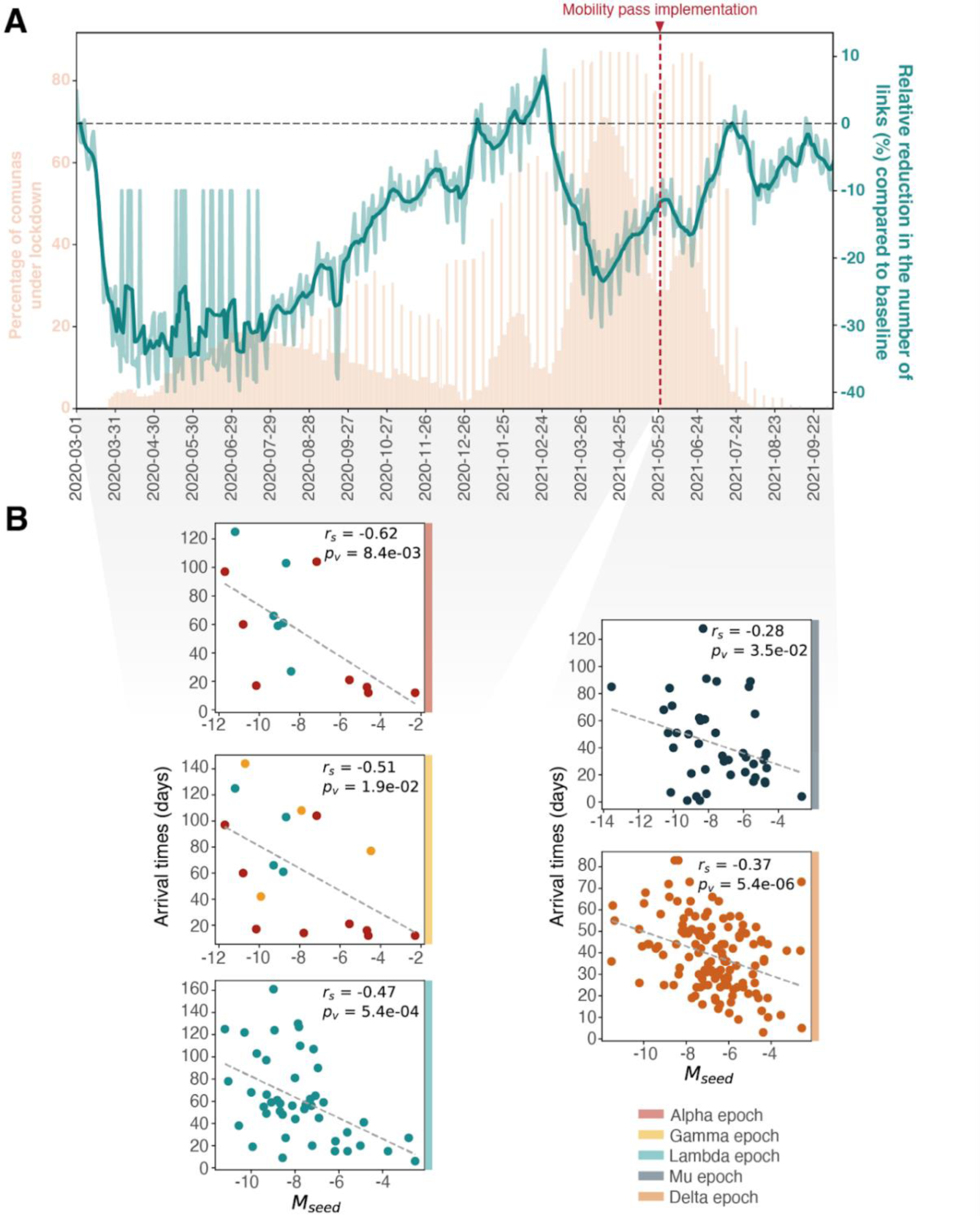
Human mobility, local NPIs and viral spread. **A)** Human mobility trends in Chile during 2020 and 2021 inferred from individual mobile phone device movements. Daily percentage of comunas under lockdown is shown for reference (light pink); this includes comunas under full lockdown or under weekend lockdown, producing the weekly spiking pattern after July 2020. The mobility metric L (see Methods) relative to a baseline level (9-15 March 2020) from before the start of the COVID-19 pandemic is shown and used to estimate the reduction of links between comunas over time. The implementation date for a Mobility Pass (May 26 2021) for fully vaccinated individuals is marked in red. **B)** Spearman correlations between the total human mobility over the epoch when variants were first introduced (shown in panel B), here referred to as Mseed, and the time to the first detection in new comunas for each TL, here referred to as arrival times.

We identified three distinct periods of human mobility within which the largest observed TLs were introduced into Chile: i) epoch 1, between January and March when large Alpha, Lambda and Gamma TLs were imported, ii) epoch 2, between May and July, when major Mu TLs were imported and iii) epoch 3, between July and September, when major Delta TLs were imported (**Fig. S6**). The arrival times of TLs to new comunas correlated with the mobility flow between the origin and destination comunas in the epoch during which the TLs were imported; this pattern is consistent for all epochs and for all variants (**Fig. 3B**). The strength of the correlation is generally greater for variants introduced during epoch 1, which predates the issuing of the mobility passes.

TLs spread rapidly across the country as a result of these seeding events, predominantly in central Chile and in large urban centres either to the south (such as Temuco and Concepción) or to the north (such as Antofagasta; **Fig. 4A**). Following importation into the country, each TL spread via short and long-range domestic seeding events into new comunas which occurred early in the invasion timeline of a TL. Some of the observed epidemic synchronicity between comunas (**Fig. S7**) is likely the result of these early seeding events taking place to locations which are both close and distant from the origin of a TL. While the timing of domestic seeding contributes to epidemic dynamics across the country, local transmission is likely affected by human mobility and mixing within specific locations which are in turn driven by different NPIs in effect before and during the occurrence of viral seeding events.

**Figure 4.**
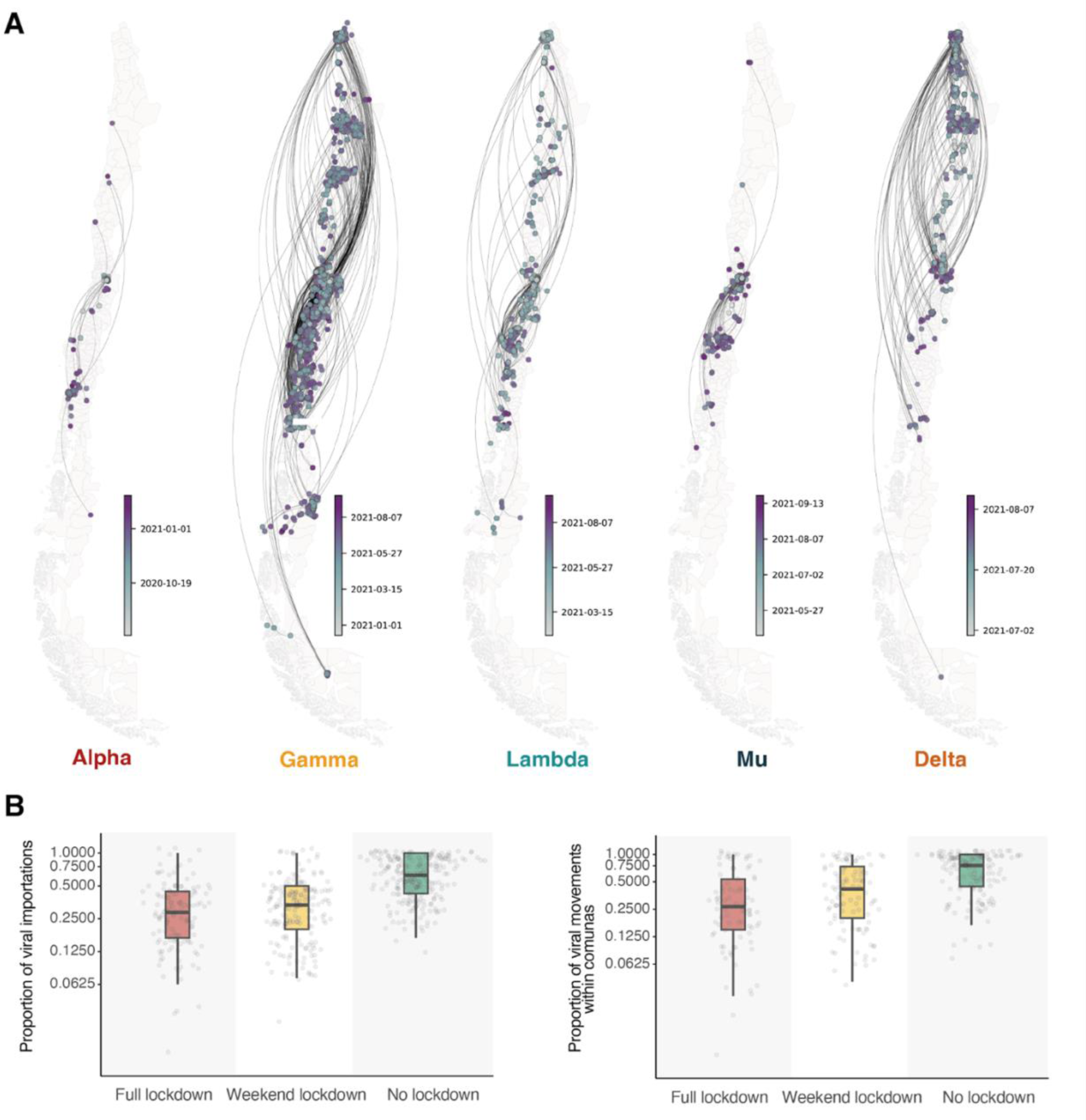
(legend). Mapping the spread of SARS-CoV-2 variants in Chile. Continuous phylogeographic reconstruction of the spread of the largest individual transmission lineage for each SARS-CoV-2 variant in Chile (Alpha TL 13, Gamma TL 35, Lambda TL 103, Mu TL 82 and Delta TL 64). Estimated median ages of tree tips and nodes are shown for each transmission lineage. **B)** Proportions of inferred viral movements by comuna for the 20 largest transmission lineages in Chile, grouped by lockdown stringency level (of the comuna where each inferred viral movement culminates). The upper panel shows inferred movements between comunas (i.e., domestic importations of viral lineages from other comunas) and the lower panel shows inferred viral movements within comunas (i.e., viral movements that start and end in the same comuna, interpreted as localised viral transmissions).

### Targeted non-pharmaceutical interventions and their effects on the spatial dynamics and persistence of SARS-CoV-2 lineages

An additional important factor in the unfolding of these epidemic waves are the highly localised NPIs implemented during 2021 following the national *Paso a Paso* plan. Despite the decentralised authority on their implementation, the stringency of these NPIs tended to be synchronised across the country (more strongly amongst comunas in the same region) and followed national epidemiological COVID-19 trends (**Fig. S8**). This created a scenario in which viral movements could occur between comunas under different lockdown stringency tiers. We hypothesise that movements within comunas should decrease with higher stringency lockdown tiers, which might also confer a protective effect on other comunas due to fewer viral imports.

To test this, we implement a negative binomial model to estimate whether higher lockdown stringency levels (n = 3) is associated with fewer inferred viral movements (modelling details can be found in *Methods*). Our results show that lockdown stringency is significantly negatively correlated with the inferred number of viral movements after accounting for new reported cases, both within comunas (χ^2^ = 7.95, *p* = 0.02) and between comunas (χ^2^ = 23.24, *p* < 0.001). Compared to comunas under full lockdown, comunas under no lockdown receive a greater number of inferred viral movements from other comunas (IRRbetween_comunas = 1.16, 95% CI: 1.06 - 1.26); comunas under weekend lockdown receive approximately the same number of viral movements from other comunas (IRRbetween_comunas = 0.97, 95%, CI: 0.88 - 1.08). These inferred viral movements are dependent on and limited by the probability of new imports generating cases that can be detected through genomic surveillance and the genomic surveillance intensity across the country (i.e., the likelihood of detecting a TL given the sampling process that produces the genomic data).

The effects of lockdown stringency on inferred viral movements within the same comuna exhibit a similar pattern: a limited and non-significant reduction is observed for comunas under no lockdown compared to comunas under full lockdown (IRR*within_comunas* = 1.05, 95% CI: 0.98 - 1.14) and comunas under weekend lockdowns show no significant differences compared to comunas under full lockdown (IRR*within_comunas* = 0.95, 95%, CI: 0.88 - 1.04). The magnitudes of these differences are small due in part to the large variation in the proportion of viral movements under each lockdown tier: essentially large numbers of viral movements are inferred for some comunas, while for others only rare and sparse viral importations are inferred (**Fig. 4B**).

A separate factor that could affect viral lineage movements in Chile is the implementation of the mobility pass for fully vaccinated individuals, to whom official lockdown restrictions did not apply. We expanded the previous model to account for the times before and after the implementation of the mobility pass on May 26. No evidence was found that the implementation of the mobility pass was associated with significant changes in the number of viral movements within comunas (χ^2^ = 1.04, *p* = 0.31), but we find a near two-fold increase (IRRbetween_comunas_mobility_pass = 1.73, 95% CI = 1.42 - 2.09) in the numbers of viral movements between comunas (χ^2^ = 29.87, *p* < 0.001).

Lineage size distributions have been shown to vary due to importation times, such that earlier importations lead to larger transmission lineages, and these larger lineages persist for longer after implementations of NPIs (30). We measure persistence using an *ad hoc* analysis of the Bayesian posterior tree distribution for the largest Gamma lineage (35, see *Methods*). We find that the proportion of persisting branches per comuna decays over time across all lockdown tiers (**Fig. S9**). The rates at which persisting branches decay over weeks vary when assessed by a fitted exponential curve (44.85%, 22.72% and 19.74% weekly reduction for comunas under full, weekend and no lockdown respectively) but wide credible intervals for all cases limit the interpretability of these differences between lockdown tiers. Future work should assess the location- and context-specific factors driving transmission lineage decay rates, including sampling intensities, geography, climate, and social interactions.

## Discussion

Our results provide an interdisciplinary evaluation of the utility of airport genomic surveillance and the role of targeted interventions in detecting and predicting viral spread of multiple viral variants. Airport and community genomic surveillance in Chile presented an opportunity to investigate the context-specific spatial dynamics of SARS-CoV-2 variants. We find that new viral importations (as inferred from genomic data) are correlated with estimates of the expected intensity of viral importations. These measurements use independent data sources, with the former relying on phylogenetically-inferred viral importations and the latter combining the flows of passengers, prevalence of the variant in the source country and the number of cases. While this has been demonstrated in other settings, previous studies have typically focussed on one viral variant at a time (1, 4, 30, 33). Incorporating the changing landscape of variant prevalence across South America revealed not only the importance of air travel but also that of land based transport from neighbouring countries, especially Chile’s large shared border with Argentina, which facilitated direct viral importations into the community, as observed for the Gamma variant. A previous study has shown the importance of land-based transport in Jordan and the Middle East (13). Airport surveillance appeared to be effective in detecting lineages at the airport and we found some evidence that it prevented these lineages from further circulating in the community (**Fig. 2D**). The earliest location of detection of a new importation does not necessarily cluster geographically with its possible source, likely due to limited spatiotemporal resolution of genomic sampling and surveillance.

The frequencies of SARS-CoV-2 variants during late 2020 and 2021 provide important context to our findings; in some regions, the pandemic was characterised by a sequence of genetic sweeps during which new variants displaced the circulating viral genetic lineages. In North America and Europe this process started with Alpha (4, 34–37) which was subsequently replaced by Delta (1, 38–41) and ultimately Omicron (33, 42–45). Similarly, in South Africa epidemic waves were driven by emerging and imported VOCs; starting with Beta, followed by Delta and finally Omicron (6, 7). Given the high connectivity of various South American countries with European countries and US states (18, 31, 46, 47), SARS-CoV-2 lineages that were dominant in the northern hemisphere heavily influenced the viral lineage composition of the region during the early epidemic waves (48–57). Nonetheless, locally emerging variants played an important role in the viral dynamics of the continent, as exemplified by the emergence and spread of Gamma (5, 58). A more extensive quantification of the differences in importation and transmission dynamics in the region could bring the possibility of predicting which variants would become dominant in a country given the regional and global context; this would also require the characterisation of both the unique immunity landscape of regions and the antigenic characteristics of novel variants.

Human mobility patterns within the country and the implementation of highly targeted NPIs played a role in the domestic spread of viral lineages within Chile. The time delay between the introduction of a new lineage in Chile and the local introduction of the lineage in a different comuna within Chile is negatively correlated with the intensity of local human movements from the source; this observation sets the basis for a probabilistic measure of viral invasion during the early stages of an epidemic as more remote comunas, which are less central in the country’s mobility network, would expect a greater delay between the epidemic taking off in central locations of the country and their own epidemic seeding.

Links between human mobility and viral seeding may also have consequences for the efficacy of NPIs, such as the highly localised comuna-level lockdowns. The reactive nature of the implementation of higher lockdown stringency levels likely resulted in a synchronised pattern of NPI stringency across the country (with strong regional synchronicity, **Fig. S8**). Viral genomic data suggests that the patterns of lineage movements across comunas show some limited differences dependent on lockdown stringency, but the limited spatiotemporal coverage from genomic surveillance poses a considerable limitation to the interpretation of these differences. Furthermore, the observed decay in the proportion of persisting lineages across all lockdown tiers follows patterns observed in other contexts (30) and it is therefore unclear the extent to which these are attributable to the NPIs themselves. A detailed analysis on the effects of these stringency levels should be paired with the quantification of the mobility changes produced by NPIs to fully understand the causal relationship of lockdowns, changes in human behaviour and the infection dynamics.

Our study has several limitations. First, after the emergence of Alpha, Gamma, and Beta, Chile implemented and increased their airport and community surveillance during 2021. This meant that the proportion of cases sequenced increased which was visible especially during the declining Delta wave at the end of 2021 (**Fig. S2**). The significantly larger number of inferred Delta introductions therefore needs to be interpreted with caution. We did however also estimate an increase of Delta EII during that time. Secondly, our EII calculations are biassed due to variable case reporting across space and time. Previous work has proposed that sequencing ∼5% of all cases allows the detection of a new viral lineage with a detection probability >80%, and that the sequencing intensities displayed by lower-middle and upper-middle income countries (as is the case for many cases in South America) allow for the estimation of a lineage prevalence with a small margin of error (59). It is unclear whether lower levels of sequencing observed for certain countries resulted in biases in genomic prevalence estimates (i.e. the true prevalence of a variant could be either higher or lower than expected), which adds further uncertainty to our EII estimates. Thirdly, the mobile phone data represent roughly one quarter of the population of Chile and it has been shown that they overrepresent urban areas and higher income groups. Accounting and adjusting for these biases will be an important area of future work. Fourth, following an importation, viral lineages spread in the country widely and circulated more intensely in key regions in north and central Chile. While continuous phylogeographic analyses are prone to sampling biases (60, 61), the distribution and circulation of these lineages around important urban areas and locations attractive for tourists suggests that viral movements in the country follow from human movements, as has been reported previously (1, 4, 9, 30, 31, 62, 63). The invasion process appears to be explained by the connectivity between comuna pairs, making human movement estimates from mobile phone usage an important predictor of arrival times of a new viral lineage into different comunas. Nonetheless, the effects of the heterogeneous NPI landscape in the country during the study period are also meaningful and likely represent an atypical mobility regime for Chile. A full description of the link between lockdown tiers and the true changes in human movement patterns (i.e., compliance with NPIs) is required to clarify the link between viral movements, human mobility and NPIs.

Coordinated genomic surveillance and the use of human mobility data can aid in the monitoring and prediction of viral spread during a large-scale national epidemic of directly transmitted pathogens. The weight of each of these data streams in the inference of pathogen lineage dynamics is an important question to address, as scaling up genomic pathogen surveillance can be costly, and data on human movements can be unavailable, or sensitive when not aggregated appropriately. Nonetheless, we find that surrogate measures such as EIIs correlate with inferred introductions, although EIIs require the collection and accessibility of epidemiological (and sometimes genomic) data from other countries. Identifying which locations act as key sources of viral importations can help to prioritise surveillance (64). Furthermore, our findings on the effect of human movement on the arrival times of viral lineages to new locations and the mitigating effects of mobility restrictions at targeted spatial scales can inform consideration of NPI packages that have a minimal effect on human freedom of movement whilst maintaining a high efficacy in epidemic control.

## Data Availability

Publicly available input data (epidemiological, flight data, land border crossings data) and scripts used to generate the analyses and plots of this manuscript are available in GitHub at https://github.com/BernardoGG/SARS2_Chile.git. The authors would like to thank and acknowledge all data contributors, i.e. the Authors and their Originating laboratories responsible for obtaining the specimens, and their Submitting laboratories for generating the genetic sequence and metadata and sharing via the GISAID Initiative. A complete list of the Authors and Originating laboratories can be found at the aforementioned public GitHub repository.

https://github.com/BernardoGG/SARS2_Chile

## Acknowledgements and funding sources

M.U.G.K. acknowledges funding from The Rockefeller Foundation, Google.org, the Oxford Martin School Pandemic Genomics programme (also O.G.P. and B.G.), European Union Horizon 2020 MOOD (#874850) (also V.C., R.P.D.I., B.G. and R.E-P.) and E4Warning (101086640) (also S.B.M.) projects, the John Fell Fund, a Branco Weiss Fellowship and Wellcome Trust grants 225288/Z/22/Z and 226052/Z/22/Z. L.F. and the authors thank the funding and support of Telefónica/Movistar Chile and CISCO Chile. This research was supported by FONDECYT Grant N°1221315 (to L.F.). LF also acknowledges financial support from the Lagrange Project of the Institute for Scientific Interchange Foundation (ISI Foundation), funded by Fondazione Cassa di Risparmio di Torino (Fondazione CRT). The views expressed are those of the authors and not necessarily those of the European Commission or any other funder.

## Author contributions

B.G., J.L.-H.T., M.U.G.K., V.C., L.F., G.P. and M.M. conceived and planned the research. A.D., R.P., S.A., and J.F. contributed an extension of the genomic and economic data, and pre-processed/cleaned the datasets. R.A. contributed socio-economic data. B.G., J.L.-H.T., M.M., R.E-P., R.D.I., G.P., and L.F. analysed the data. B.G., and M.U.G.K. wrote the first draft of the manuscript. All authors subsequently edited, read, and approved the manuscript.

## Materials and Methods

### SARS-CoV-2 sampling from community and airport surveillance in Chile

Chile implemented a systematic genomic surveillance programme in April 2021. During epidemiological week (epiweek) 47 of 2020 (2020-11-21 to 2020-11-27), randomisation of community samples for the early identification of variants of interest was implemented through PCR testing of variant-associated mutations (65, 66). This strategy considers random, representative sampling without incorporating clinical or epidemiological criteria to estimate the prevalence of SARS-CoV-2 variants and lineages circulating in Chile, using a weekly sample size based on the number of new cases registered the previous week. During epiweek 5 of 2022 (2022-01-30 to 2022-02-05), this strategy was updated for sampling from Arturo Merino Benítez International Airport (in this work referred to as the Santiago International Airport or SCL for short) and the laboratory requirements for submitting samples to the Instituto de Salud Publica (ISP) for genomic sequencing of SARS-CoV-2 cases (67). The strategy provided specific details regarding the sample size, which was based on the number of cases in travellers during the previous epiweek (with a novel variant prevalence set at 0.50%, or 1/200, and considering a 95% confidence level; (68). This approach aimed to identify novel variants entering the country and estimate the incidence of circulating variants in travellers.

### Data from COVID-19 epidemics in Chile and connected countries

Epidemiological, genomic and human mobility data was collected between November 2020 (which includes the earliest estimated ancestor of a transmission lineage in Chile with a TPMRCA on 2020-11-01; TPMRCA referring to the time of the parent node of the earliest tree node inferred to have occurred in Chile, followed by local viral transmission inside the country) and October 2021 (which includes the latest sample collection date for our study time period on 2021-10-12). This time span is henceforth referred to as the ‘study period’.

The monthly numbers of air travellers entering Chile during the study period were collected from the National Civil Aviation Agency and the National Ministry of Transportation and Telecommunications (Junta Aeronáutica Civil, Ministerio de Transportes y Telecomunicaciones) at http://www.jac.gob.cl/estadisticas/informes-estadisticos-mensuales-del-trafico-aereo/ (retrieved on 03-08- 2022). This data includes the source city from which passengers arrived into Chile, as well as the airport to which they arrived. Given that 99.24% of returning travellers entered the country through the Santiago de Chile International Airport (SCL) and that all other international airports only served travellers from 8 countries (Argentina, Colombia, Peru, Paraguay, Venezuela, Bolivia, Haiti and Uruguay) out of 23 countries where travellers arrived from, data from airports outside of SCL was not considered for further analyses as their overall contribution was negligible. Furthermore, genomic data from airport surveillance was only collected at SCL, making the use of this data more comparable to our phylogenetic inferences. Given that counts for incoming air travellers was only available on a monthly basis, the total number of passengers per month was divided equally across the weeks that make up each month to obtain weekly estimates. This transformation assumes equal numbers of passengers entering every week from each origin city, which is justified by the periodic frequencies of airline scheduling practices (69). The monthly number of individuals entering the country through land border crossings was obtained through an Information Transparency request to the government of Chile data portal (https://datos.gob.cl). This data included the name of the specific border crossing station at which the numbers were recorded, from which we can identify the source country from where travellers entered and the comuna in Chile where they first arrived. We also divided the numbers of travellers equally across weeks in any given month for convenience, even if the periodic flight schedule assumption is unlikely to apply to land border crossing data.

Anonymised, individually reported COVID-19 cases from Chile were provided by the ISP of Chile. For each case, the sample collection date and comuna (lowest administrative level in Chile equivalent to adm3) of residence of the patient are recorded. Cases are aggregated daily and by comuna to produce epidemiological time series for the country. During the study period, Chile established a unique decentralised system for non-pharmaceutical interventions (NPIs) where individual comunas had autonomous authority to place their area under one of three tiers of stringency on limitations to human movements and gatherings (**Supporting Text**). Daily records for the NPI stringency tier for each comuna were used to estimate the total population under lockdown on any given day by multiplying the number of comunas under lockdown by their respective population sizes (provided within the same data files, obtained through the public COVID-19 GitHub repository of the Government of Chile, available at https://github.com/MinCiencia/Datos-COVID19/tree/master/output/producto24). Population sizes and aggregated case counts per country for locations from which incoming travellers were recorded into Chile during the study period were obtained from the Our World in Data COVID-19 dashboard at https://ourworldindata.org/covid-cases (retrieved on 05-12-2021; (70)).

Genomic data from Chile was generated through two distinct genomic surveillance programmes and is described in the *SARS-CoV-2 genomic data from community and airport surveillance in Chile* section. Metadata for these sequences separated by surveillance programme was provided by the ISP, including an individual sequence ID which matches the ID assigned to viral genomes when uploaded to GISAID (71). Genome sequences from Chile were downloaded from GISAID (retrieved on 2022-06-30) and filtered by matching individual IDs with the metadata provided by the ISP.

#### Generation of a background genomic data set utilising international human mobility data

Global genome datasets assigned to each variant under investigation (Alpha, Gamma, Lambda, Mu and Delta) were downloaded jointly with the Chile sequences from the aforementioned GISAID dataset. To gain an overview for the global introductory events into Chile, we curated a bespoke global dataset using openly available human mobility data. For each dataset, all global sequences were sorted by their location and their relative contribution to all international human movements into Chile. The top 5 countries with the highest relative international movements into Chile were included in our dataset, in addition to the top 7 countries outside of South America with direct flights to Chile. All datasets were then subsampled uniformly across time per epidemiological week over the study period. This resulted in datasets with an approximate 1:1:1 ratio of sequences from Chile versus sequences from countries with the highest relative mobility versus countries with the most direct flights from outside of South America: 609 sequences for Alpha, 4826 sequences for Gamma, 3164 sequences for Lambda, 1720 sequences for Mu, and 6983 sequences for Delta.

#### Extraction of mobility estimates from mobile phone network data

We analysed up to 3.5 million individual daily trajectories (which correspond approximately to 24% of the Chilean mobile phone subscription market share) from a total of 6.5 million Telefonica mobile phone users. We then aggregate individual trajectories into mobility indicators, selecting three main mobility aggregation methods, and we proceed by evaluating the interplay of the mobility process, with the spatial invasion. The first aggregating method counts the number of displacements between any two consecutive eXtended Detail Records (XDRs) made by a user, and it accounts for the full trajectory of each individual (D). The second coupling matrix connects the residence location of each user to all their visited locations, with a coupling force that is proportional to the number of XDRs made in each location (L). The third coupling matrix (L1) connects the residence location of each user to all their visited locations, with a coupling force that is proportional to the time spent in each location (72). We quantified the associated coupling matrices (D, L, L1) for each month during the study period in Chile at the comuna level.

#### Phylogenetic inference of SARS-CoV-2 viral importations into Chile

Variant-specific genomic data sets were aligned to the Hu-1 reference genome (73, 74) using Minimap2 (75), and a maximum likelihood (ML) phylogenetic tree was estimated for each alignment using IQtree (76) under a GTR substitution model and by modelling the evolutionary rate variation across sites using a Gamma distribution with four site categories. The temporal signal for each phylogeny was evaluated by estimating the regression between root-to-tip lengths and sample collection dates using TempEst (77), after establishing the most likely root for each phylogeny by a minimisation of residuals from each regression. Sequences for which the residuals were estimated to fall more than two standard deviations from the mean were removed, and the ML tree re-estimated under the same conditions described above. For these phylogenies, polytomies were not randomly resolved but rather maintained in the trees.

Branch lengths for the inferred ML trees were re-scaled to fit a temporal timescale using TreeTime (78). For all phylogenies, a fixed evolutionary rate of 7.4×10-4 subst/site/year was used for these analyses, following the empirical findings by Ghafari et al (79). Given that individual analyses were performed per variant, the use of a fixed evolutionary rate allows us to ignore the likely inflated evolutionary rate observed prior to the emergence of individual variants (80) while also minimising potential comparability issues among data sets due to the increased rate variation amongst lineages due to the different data set sizes and sampling periods which can obscure the temporal signal (81).

Time-scaled ML phylogenies were further analysed using an alignment-free likelihood estimation method implemented in BEAST v1.10.5 (pre-release; commit:d1a45) commonly known as Thorney BEAST, and described in further detail in (30). The time-scaled ML phylogenies were used as starting trees, while the previously described polytomic ML phylogenies were used as data trees; Thorney BEAST samples from different node heights and resolutions for polytomies to generate a posterior sample where branch length (in genetic terms; i.e., number of mutations) likelihood estimates are calculated as a function of a Poisson distribution with mean equal to the evolutionary rate multiplied by the branch length (in time units; (1)). Individual MCMC chains for each data set were run for 100 million steps with a burn-in of approximately 10% of the initial steps. All runs had a fixed evolutionary rate of 7.4×10-4 subst/site/year and a non-parametric Skygrid tree prior (82) with breakpoints every two weeks of the sampling period for the specific variant being analysed. Trees were sampled every 5000 steps, and convergence of all parameters was evaluated using Tracer (83) and defined as parameter effective sample sizes (ESS) higher than 200.

The resulting empirical tree distributions were reanalysed using BEAST 1.10.4 to estimate the number of importations into Chile using a discrete trait analysis (DTA) phylogeographic approach. Sequences collected from outside of Chile were labelled as “non-Chile”; sequences collected within Chile were further identified as “airport” or “community” sequences according to whether they were collected through airport surveillance of returning travellers or community surveillance inside Chile, respectively. Transitions between discrete states were estimated using an asymmetric model, and individual counts of transitions between states were estimated by stochastic mapping in the form of Markov jumps and rewards (84, 85). These DTA analyses were run for 10 millions MCMC steps and sampled every 10 000 steps. Summary maximum clade credibility (MCC) trees were generated for all data sets using TreeAnnotator.

#### Phylogeographic reconstruction of the domestic spread of viral transmission lineages in the country

From the discrete phylogeographic analyses, individual importations were identified following the rationale used by du Plessis et al (30): subtrees that descend from a node inferred to have occurred inside Chile, which in turn descends from a node inferred to have occurred outside Chile. These subtrees, referred to as transmission lineages (TLs, (30)), were extracted using Fertree (https://github.com/jtmccr1/fertree). We analyse the 20 largest TLs (Alpha, n = 1; Gamma, n = 2; Lambda, n = 5; Mu, n = 5; Delta, n = 7) which correspond to those with sizes greater than the 93rd percentile of all identified lineages within the country (> 61 tips). These transmission lineages range in size from the largest being Gamma TL 35 (n = 1827) for Gamma (corresponding to the Chile-specific PANGO designated Gamma sublineage N.4) to the smallest being Mu TL 61 (n = 61) for Mu. We extracted these larger transmission lineages from the summary MCC trees and performed individual continuous phylogeographic analyses for each to map their spread across comunas within Chile.

Sequence metadata was available for sequences up to the comuna (adm3) level, but no individual georeferenced coordinates were collected to ensure patient privacy. For each individual sequence where the comuna sampling location was known, we randomly assigned a grid cell with sample weights proportional to the population density within the comuna obtained from WorldPop, at a resolution of 1km^2^ grid (https://hub.worldpop.org/geodata/summary?id=44918). Sequences assigned to the same grid cell were further given random coordinates within the grid with uniform sampling to ensure that all sequences have unique coordinates as required by the continuous phylogeographic model. A final check was placed to ensure that the final coordinates fell within the comuna polygon.

For the 20 extracted subtrees, we pruned all sequences not collected in Chile and assigned the aforementioned coordinates to the remaining tips. We then used BEAST to estimate the continuous diffusion of the TLs using a relaxed random walk (RRW) model with a Cauchy distribution to account for among-branch dispersal velocity heterogeneity (32). MCMC chains were run by duplicate for 50 000 000 steps each, with multiple runs performed when necessary to achieve convergence. Multiple chains for each TL were combined using LogCombiner, and the summary MCC trees for each TL were generated with TreeAnnotator. Mapping the dispersal of each TL in Chile was done with custom Python scripts (https://github.com/leoferres/spatial_dynamics_covid_chile).

#### Estimated importation intensity (EII) indices from variant prevalence, case, and human travel data

To estimate the expected number of importations of each VOI/VOC into Chile over time independently from our phylogenetic inference, we compute a weekly estimated importation intensity (EII) index for every country from which incoming flights into Chile were recorded during late 2020 and 2021. Exceptions to the country-level EIIs were considered for the USA and Brazil where state-level estimates were produced instead to account for the broad geography of both countries and their heterogeneous epidemiological landscapes. Five states from the USA (Florida, New York, Texas, Georgia and California) and seven states from Brazil (São Paulo, Rio de Janeiro, Goias, Parana, Rio Grande do Sul, Bahia and Santa Catarina) were included as these are the only states in each country from which direct flights arrive to SCL.

For any variant under investigation *X* (Alpha, Gamma, Lambda, Mu, Delta) and source location *l,* the **EII** over *n* epidemiological weeks is defined as

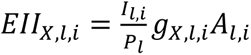

where *Il,i* is the aggregate number of new cases for epidemiological week *i* in location *l*, *Pl* is the population from location *l, gX,l,i* is the proportion of viral genome sequences (available in GISAID) that were sampled at location *l* in week *i* and that are assigned to variant *X*, and *Al,i* is the total air passenger volume coming into Chile from location *l* on week *i*. For the three neighbouring countries (Argentina, Bolivia and Peru) that share a land border (and therefore border crossings) with Chile, variation of the EII was estimated (***l*EII**) as

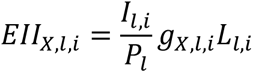

where *Ll,i* is the number of land travellers entering Chile from location *l* on week *i*. For these countries, the air-travel-based EII described previously (referred to as ***a*EII** when relevant) was combined with the ***l*EII** to estimate a global EII as

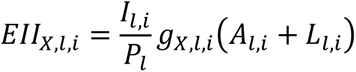

Since this formulation of the EII estimates the numbers of individuals expected to enter the country infected with variant *X*, we also derive a combined ***c*EII** from all locations from which international air travellers come to Chile:

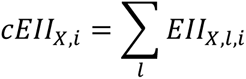

*Vector autoregressive (VAR) models to test forecasting power of EIIs on viral imports per variant* Given the weekly estimates, every EII variation for each individual viral variant *X* is concatenated into a time series covering *t* = 48 epidemiological weeks, describing the expected number of introductions per variant over time. These EII time series are compared to time series of inferred observations of viral importations from the genomic data *P*, taken as the weekly aggregate counts of phylogenetic nodes which are the most recent common ancestors of individual Chile transmission lineages. We formulate a bivariate vector autoregression model from both time series:

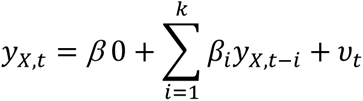

Where *y_x,t_* is a vector of both *P* and EII values at week *t* and *k* is the maximum lag, equal to 12 to represent a maximum allowed delay between expected and inferred importations of approximately three months. From this, the system of equations is:

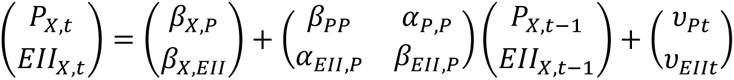

The optimum value for k is evaluated by comparing information criteria from each lag value using the vars package in R; we specifically evaluate the Akaike Information criteria (AIC), Hannan-Quinn information criteria (HQ), Schwartz criteria (SC) and Akaike’s Final Prediction Error (FPE). We take the lowest value amongst the four indicators as the indicator of the optimum lag value indicator, which in most cases corresponds to SC or AIC (**Table 1**). To test whether phylogenetically inferred importations P are forecasted by the EII estimates, we perform Granger causality (GC) tests with α=0.05. Contrary to what its name suggests, GC tests do not establish causality between the time series but rather temporally dependent correlation; we therefore refer to our findings as one time series Granger-causing the other, which is not true causality.

#### Estimation of the effects of human mobility at the comuna level on viral movements

Following a viral importation, some transmission lineages (TLs) spread domestically within Chile. The comunas where these local TLs were first detected are not a reliable measure of where the introduction likely took place due to delays between importation and detection as has been shown previously (30) (**Fig. S10** shows the numbers of ‘first detection’ events in the country and the location of land border crossings in the country; the lag between the TMRCAs and the first detection of TLs in Chile was estimated to be 9.59 ± 8.95 days). Instead, we estimate the impact of local scale human mobility on the invasion dynamics of different TLs by comparing comuna-to-comuna specific arrival times extracted from our continuous phylogeographic analysis to human movements.

For each epoch *v*, we compute the metric of mobility exchange between the source comuna *s* and location *j*, namely *M_j_^v^* as:

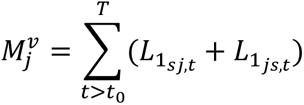

where *j* represents comunas, *t* runs from *t0*, day of first detection of the variant defined in epoch *v* in the source comuna *s*, to *T*, days to consider for the mobility aggregation, *L1sj,t* is the time spent by residents of *s* in *j* on day *t*. *Mj^v^* accounts for the total time spent respectively by users of the source location in comuna *j* and vice versa. Epochs *v* are defined as 60-day intervals starting on the first date of detection of a variant (which defines the epoch) at comuna level and are named after the variant itself. The time window of mobility aggregation *T* must be shorter than the epoch, in Fig. 4 we chose *T* = 2 weeks. For each epoch, only transmission events with both transmission start date and respective source time *t0* falling within the epoch time window are chosen. Epochs may include multiple TLs spreading at the same time. Arrival times at comunas *j* are then compared with the human mobility between comuna *j* and the mobility between *j* and source *s*.

#### Estimation of the effects of NPI stringency at the comuna level on viral movements and viral lineage persistence

We formulated simple negative binomial models to test whether the stringency of NPIs implemented at the comuna-level had a measurable effect on the estimated numbers of viral movements within a single comuna or between comunas. For every inferred viral movement (represented by a branch in the continuous phylogeographic analysis, with the ancestral node location accounting for the source comuna for that movement and the descendant node or tip location accounting for the destination comuna for that movement), we determine the lockdown stringency tier that corresponds to the estimated median age of the source and destination comunas, and aggregate all movements based on the lockdown tier of the destination comunas. The models estimate the aggregated counts of viral movements (either into each comuna or within a comuna) explained by both the lockdown stringency tier (three levels: No lockdown, weekend lockdown or full lockdown) and the numbers of new cases reported during the duration of that lockdown. We confirmed the appropriateness of a negative binomial model compared to a Poisson regression model (i.e. the assumption of conditional means not being equal to conditional variances) by performing a likelihood-ratio test between both model fit methods (DF = 1, *p* < 0.001). From the fitted model coefficients we estimate the incidence rate ratios (IRR) to quantify the effect of lockdown tier stringency on viral movement counts. We further expand these models to also account for the effect of the implementation of a mobility pass for fully vaccinated individuals on May 26, 2021; results for both model formulations are shown even if a model which doesn’t account for the mobility pass better fits the data for viral movements within comunas (LRT = 1.04, *p* = 0.31) while accounting for the mobility pass is a better fit for the data of viral movements between comunas (LRT = 29.87, *p* < 0.001).

Following the arrival of viral lineages to a new location (comuna), they could either persist and be detected within that location or become extinct. The number of newly arrived lineages and previously circulating lineages in a location can be used to estimate the proportions of persisting lineages (46, 86). We analyse the posterior tree distributions from the continuous phylogeographic analyses using *PersistenceSummarizer* (86) to estimate the number of persisting lineages within the fourteen comunas with the highest number of observed viral movements (five comunas in the Santiago Metropolitan Area: Pirque, Puente Alto, Santiago, San Jose de Maipo and San Bernardo; nine comunas from the rest of the country: Arica, Los Angeles, Requinoa, Copiapo, Iquique, Valparaiso, Antofagasta, Concepcion and Puerto Montt) under different lockdown stringency tiers. We aggregate comunas under the same lockdown tier and set the date when the tier was enacted as the ancestral time, and follow up with sequential evaluation times on every week following the implementation of the lockdown stringency tier. This way, we can estimate persistence trends in different comunas under the same lockdown stringency tier despite the fact that these were implemented at different points in time, and that stringency tiers were enacted and relaxed on multiple occasions across our study period. Full and weekend lockdowns were implemented for a maximum continuous time of 16 and 20 weeks respectively (**Fig. S11**), we therefore perform lineage persistence analyses for a maximum of 25 weeks following the implementation of a specific stringency tier. The remaining comunas beyond the fourteen mentioned above are excluded as they display few viral movements and therefore yield insufficient information regarding lineage persistence.

To estimate the rate of decay of persisting lineages under different stringency tiers, we fit an exponential model to the median number of persisting lineages over every week following the implementation of the specific tier. The model is formulated as:

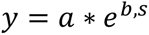

Where *y* is the median proportion of persisting lineages from Gamma TL 35 and *x* is the number of weeks since the implementation of that lockdown tier. Decay rates are estimated as model parameter *b*.

### Supporting text - Contextualising the COVID-19 and mobility situation in Chile during 2020-2021

Located on the southernmost end of the South American Pacific coast, Chile provides an interesting context to analyse the effects of human mobility and spatial connectivity in the spread of SARS-CoV-2 as it includes highly connected yet largely rural areas. The majority of air travellers enter the country through the Arturo Merino Benítez International Airport, located on the outskirts of its capital city Santiago de Chile (in this work referred to as the Santiago International Airport or SCL for short), which accounted for 99.24% of all travellers entering the country through international flights between october 2020 and december 2021 (the remainder entering through six secondary airports, as shown from publicly available data from the Junta Aeronáutica Civil, Ministerio de Transportes y Telecomunicaciones; http://www.jac.gob.cl/estadisticas/informes-estadisticos-mensuales-del-trafico-aereo/). While this single main port of entry drives registers a large portion of all incoming international travellers, the country also shares a long border with neighbouring Argentina, the longest international land border of South America, covering 5,308km in length; this border features 26 official international border crossings through which substantial numbers of travellers enter Chile, most predominantly into the regions of Atacama in the north (Paso Sico), Coquimbo (Paso Agua Negra) and Valparaíso (Paso Los Libertadores) in central Chile (including Santiago) and Los Lagos (Paso Cardenal Samoré) in the south of the country. Overall, land border passes register up to one third of all international arrivals into Chile (e.g. 652,843 travellers entered Chile from border crossings in 2021 as shown from data obtained through an Information Transparency request, see *Methods*). Internally, Chile has a generally well developed highway infrastructure and a domestic flight network, but its elongated geography along the north-south axis lends itself to the establishment of remote areas across the national territory with limited human mobility. Approximately 38.62% of the population lives within the Santiago Metropolitan Area, with an additional 15.08% of the population residing in the major urban centres of Valparaíso, Concepción, Temuco and Antofagasta (2017 census data available at https://www.bcn.cl/siit). Population densities tend to be high in the central regions of the country (1); the rest of the country is less densely populated, including large and mostly unpopulated natural parks and remote regions. The vast Atacama desert in the north and the extreme latitude of the southernmost regions of Chile might discourage the development of large human settlements but incentivise a rich tourism industry.

As with other countries, Chile suffered local COVID-19 epidemic waves throughout 2020 and 2021. The public health response to SARS-CoV-2 in Chile included several key particularities. From a surveillance perspective, the country implemented had been running genomic surveillance since 2020 and implemented a systematic genomic surveillance programme in April 2021 after the emergence of the Alpha and Gamma VOCs amongst others (2); within this broader programme, specific testing was performed on all international returning travellers at the Arturo Merino Benítez International Airport (in this work referred to as the Santiago International Airport or SCL for short). This dedicated testing programme was used not only to inform travellers about their quarantine requirements if they tested positive, but also served as a sentinel system to identify incoming viral lineages and specifically VOIs/VOCs. As non-pharmaceutical interventions (NPIs), Chile implemented a targeted scheme for partial and complete lockdowns and progressive reopening at the *comuna* level (adm3 administrative divisions; n = 346 communas) called *Paso a paso nos cuidamos* (lit. “Step by step we take care of ourselves”) in July 2020 (3). *Comunas* were placed in one of five possible lockdown tiers (called ‘steps’, lit. *Pasos*), in order of stringency these are ‘*Cuarentena*’ (Quarantine), ‘*Transición*’ (Transition), ‘*Preparación*’ (Preparation), ‘*Apertura inicial*’ (Initial opening) and ‘*Apertura avanzada*’ (Advanced opening). Limitations within these tiers concerned, amongst other things, the requirement of special permits to enter/leave the comunas or to circulate freely. In practice the different levels of stringency resulted in the following mobility characteristics:

1. The **Quarantine step** limited human mobility throughout the entire week
2. The **Transition step** limited human mobility only on weekends when the stringency of the higher tier (Quarantine) was maintained.
3. The **Preparation** and **Initial opening steps** share similar mobility regimes, with no restrictions on free circulation in the comuna and the main differences between them concerning specific types of activities permitted and recommendations on numbers of people for gatherings.
4. The **Advanced opening** step results in scenarios with no restrictions.

As such, we re-code the NPI stringency levels into three tiers: ***(1) full lockdown*** (includes comunas in Quarantine step), ***(2) weekend lockdown*** (includes comunas in Transition step) and ***(3) no lockdown*** (includes comunas in the Preparation and Initial opening steps). The scheme provided some degree of autonomy to individual *comunas* and allowed them to decide when to increase or reduce the lockdown stringency level. This resulted in a highly heterogeneous landscape of the timing and intensity of non-pharmaceutical interventions across the country; the effects of such a spatially heterogeneous implementation of interventions on viral transmission remain unclear.

**Figure S1.**
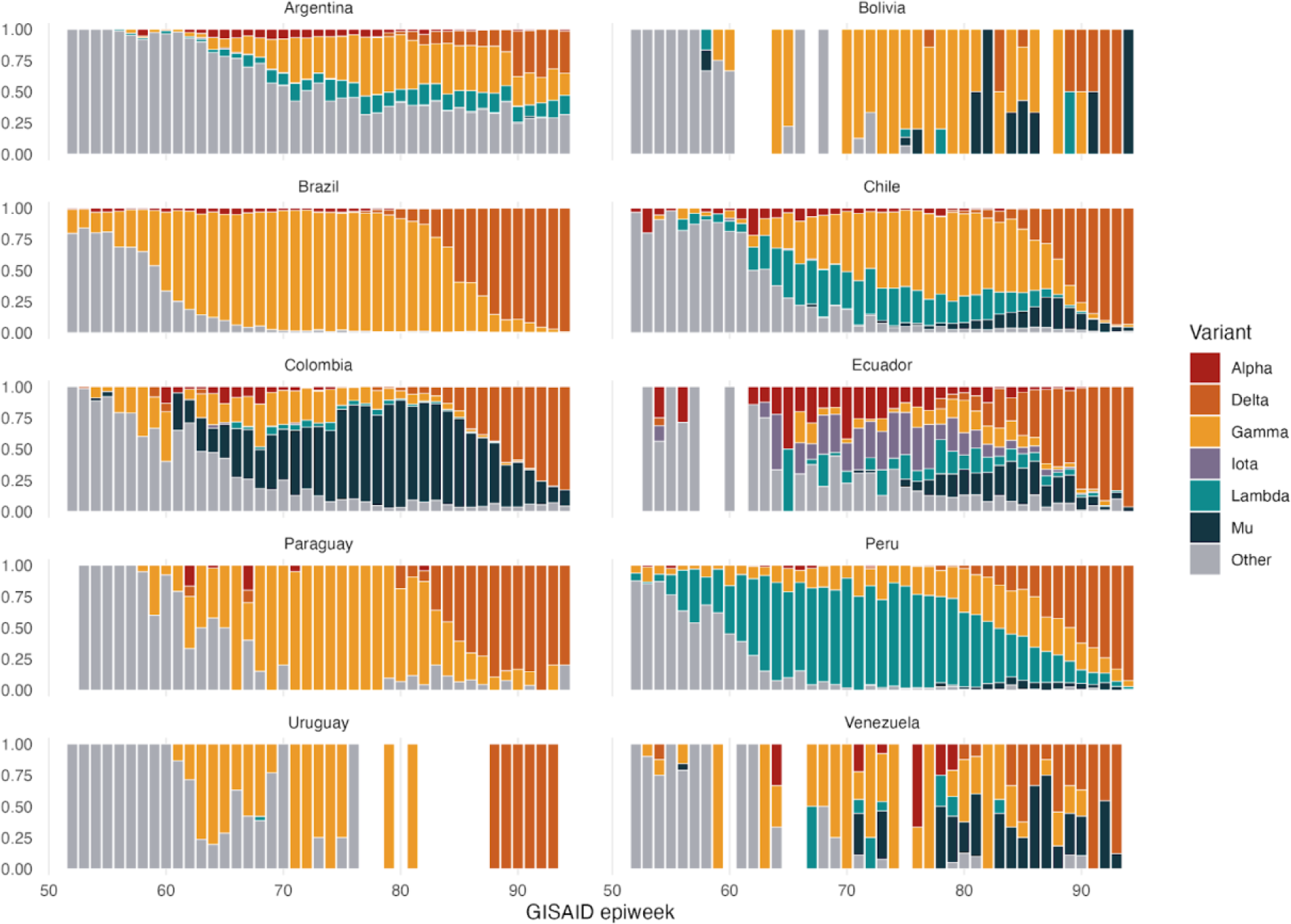
Proportion of SARS-CoV-2 variants detected in South American countries. The proportion of sequences corresponding to each VOI/VOC per epidemic week (as annotated on GISAID) is shown per country, relative to the total number of sequences generated by each country over that two-week period.

**Figure S2.**
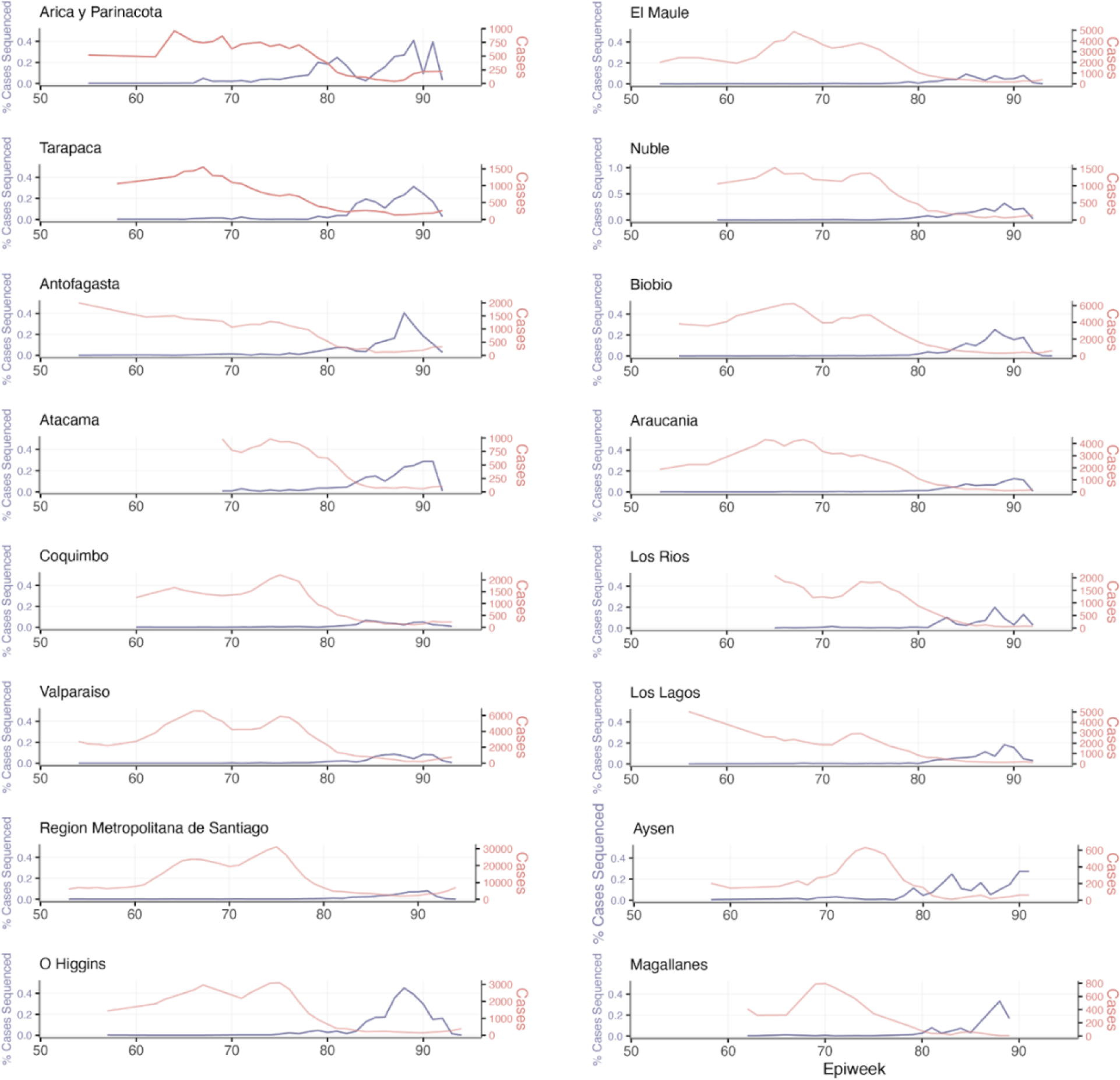
Epidemic trends and sequencing intensity by geographic region in Chile. Numbers of cases and percentage of cases sequences in 16 geographic regions of Chile per week over 2021.

**Figure S3.**
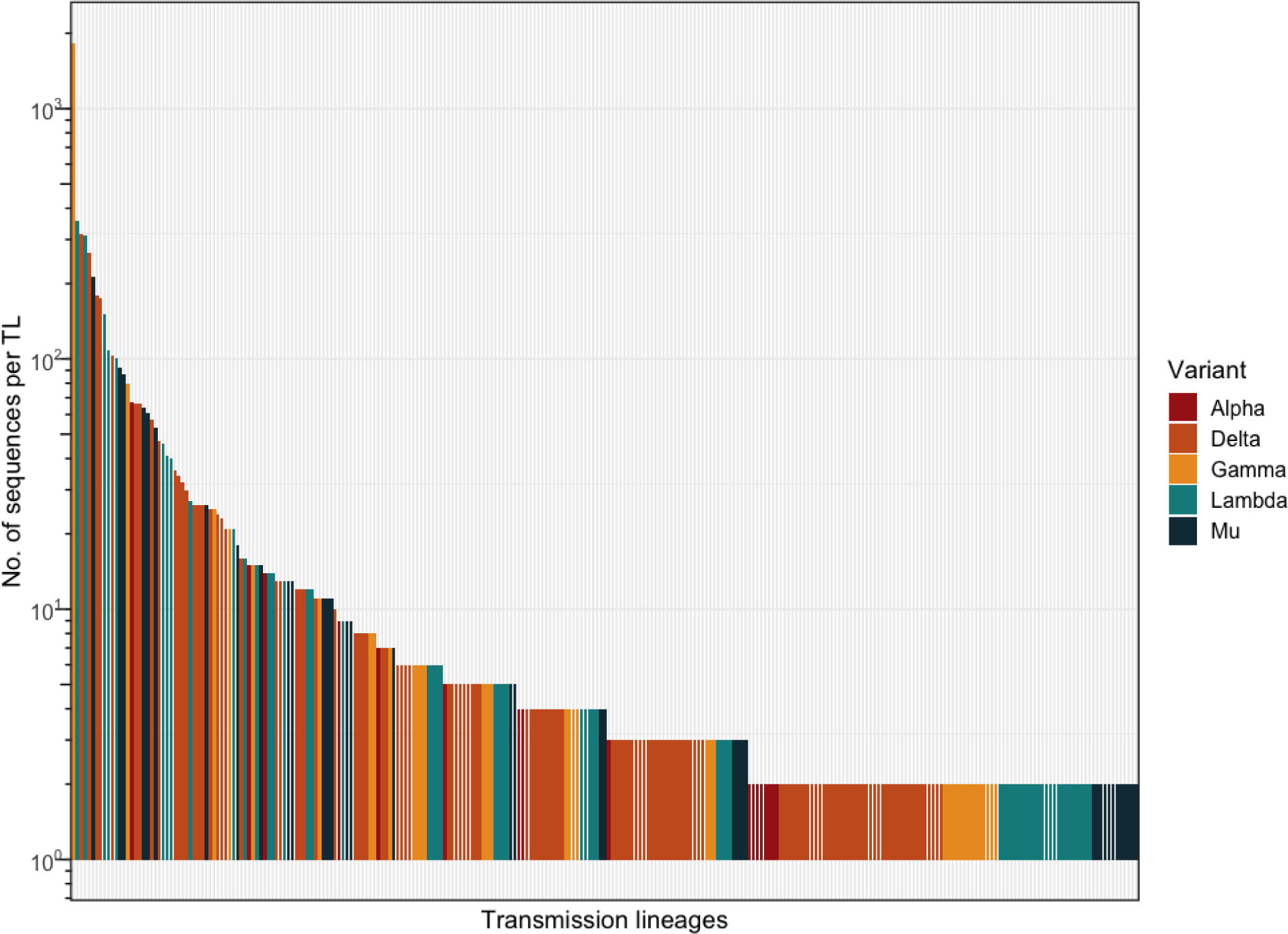
Distribution of transmission lineage sizes. Distribution of the number of sequences in each transmission lineage inferred from genomic data (includes singleton, i.e. transmission lineages with n = 1 taxa) in Chile coloured by variant. Number of taxa shown on a logarithmic scale.

**Figure S4.**
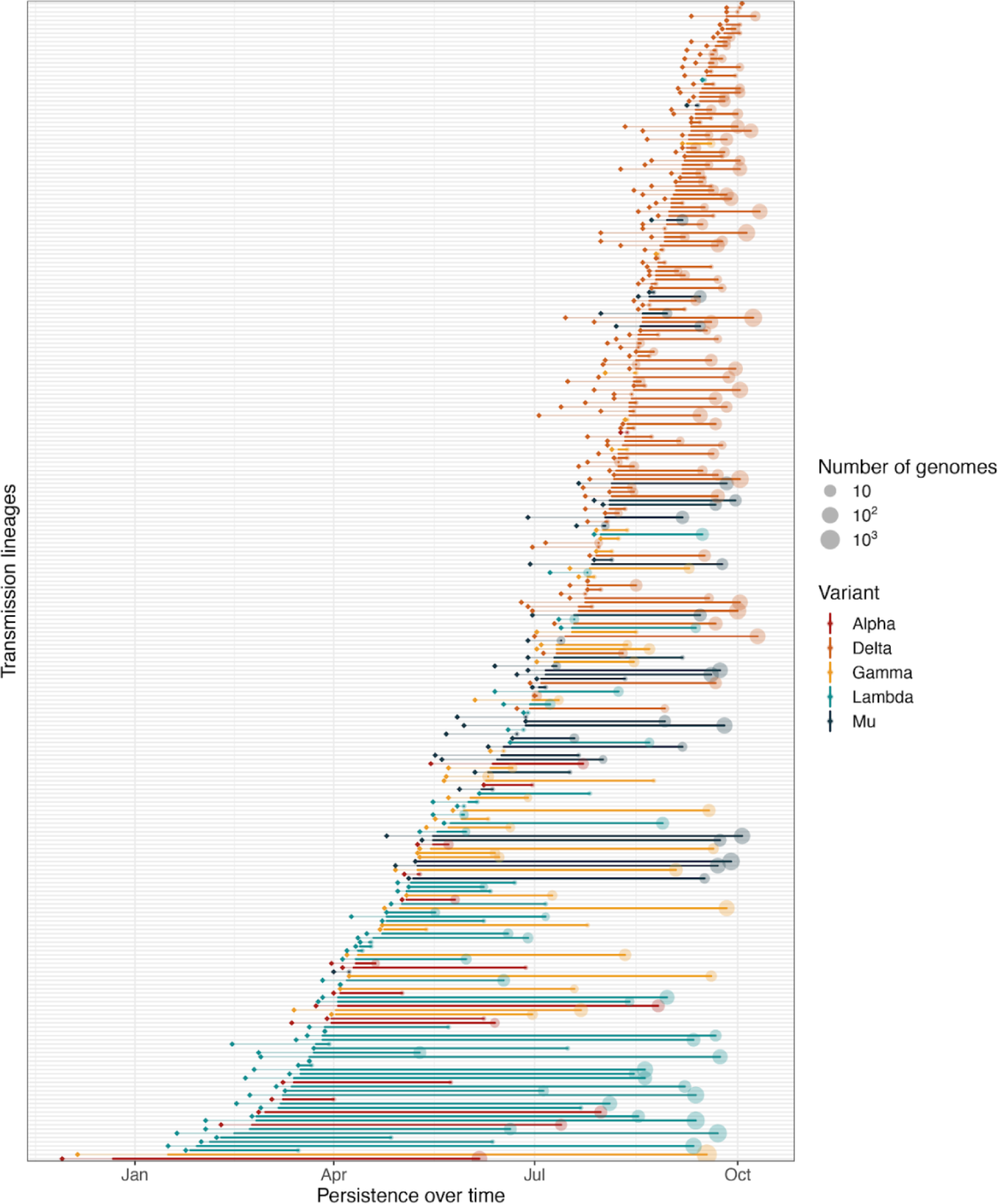
Transmission lineage TMRCAs and persistence over time. Rows show individual transmission lineages (singletons excluded, i.e. transmission lineages with n = 1 taxa) plotted over the time when they were detected and circulating in Chile. Solid lines show the detection period (time lapse between the collection date of the earliest sequence, referred to as ‘first detection’, and the collection date of the most recent sequence, referred to as ‘last detection’). Faded lines show the detection lag period (time lapse between the inferred transmission lineage TMRCA, shown as diamonds, and the date of first detection). Size of circles on the date of last detection shows the number of sequences for that transmission lineage.

**Figure S5.**
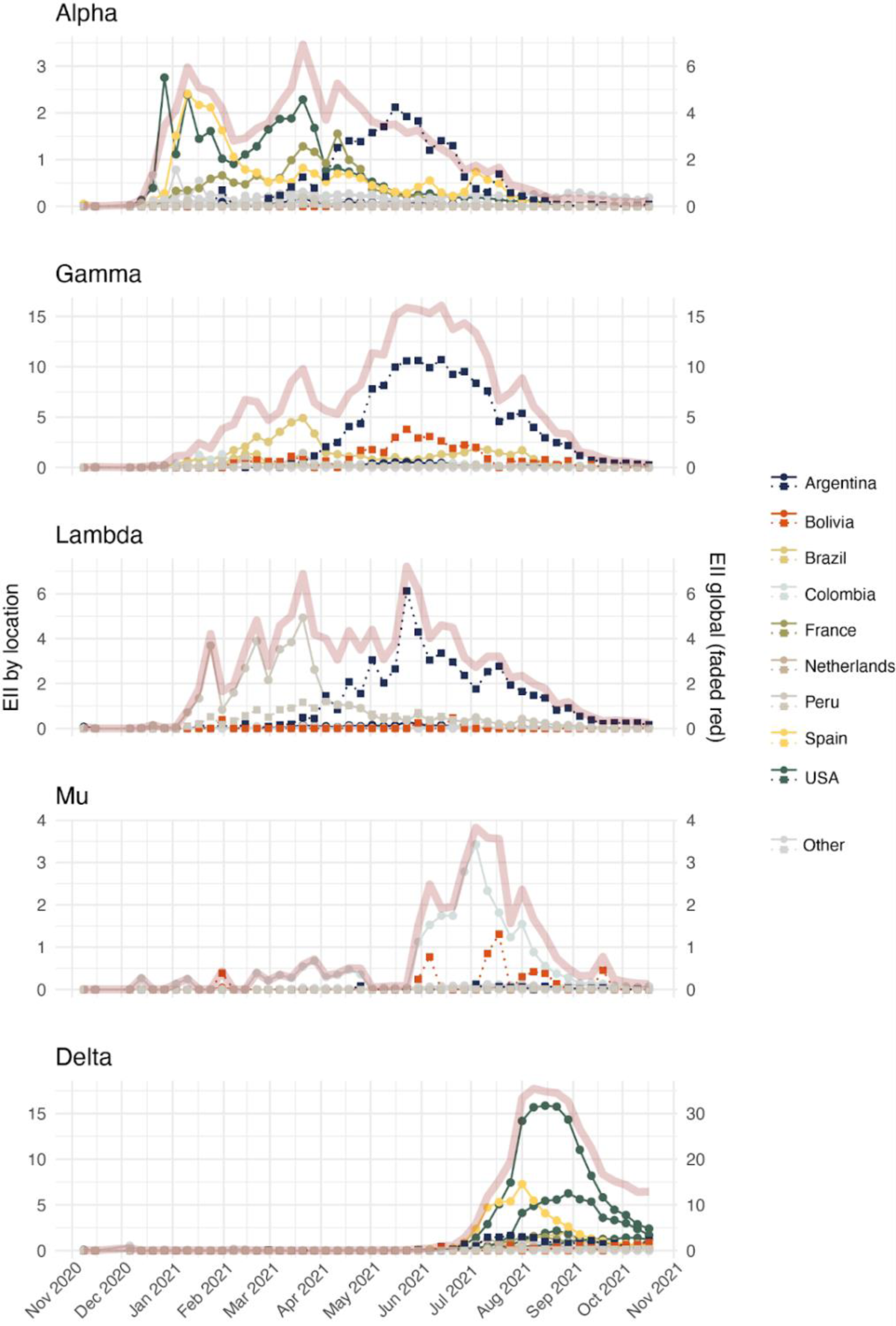
Estimated importation intensity (EII) indices over time. Time series for the weekly estimated importation intensity (EII) indices from selected states and countries related to Figure 2C. Solid lines with circles show EIIs based on air travel volume (aEII) and dashed lines with squares show EIIs based on land border crossings (lEII). Broad red lines show the combined global EII.

**Figure S6.**
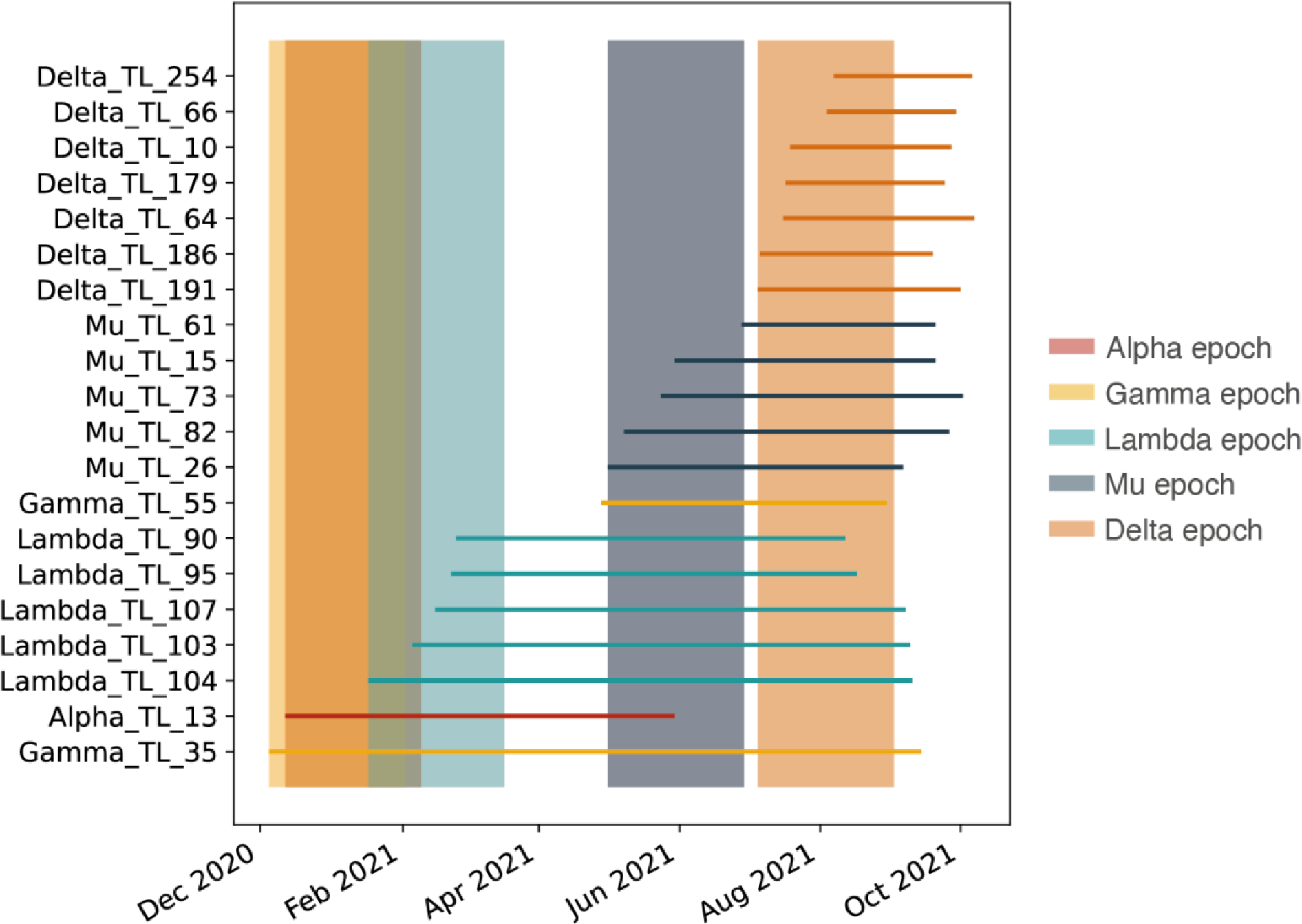
Time of arrival epochs. Importation dates of the 20 largest SARS-CoV-2 transmission lineages in Chile coloured by variant. Coloured bars show the epoch over which mobility estimates were used to evaluate their effect on viral spread, depending on the dates of the first importation of each variant.

**Figure S7.**
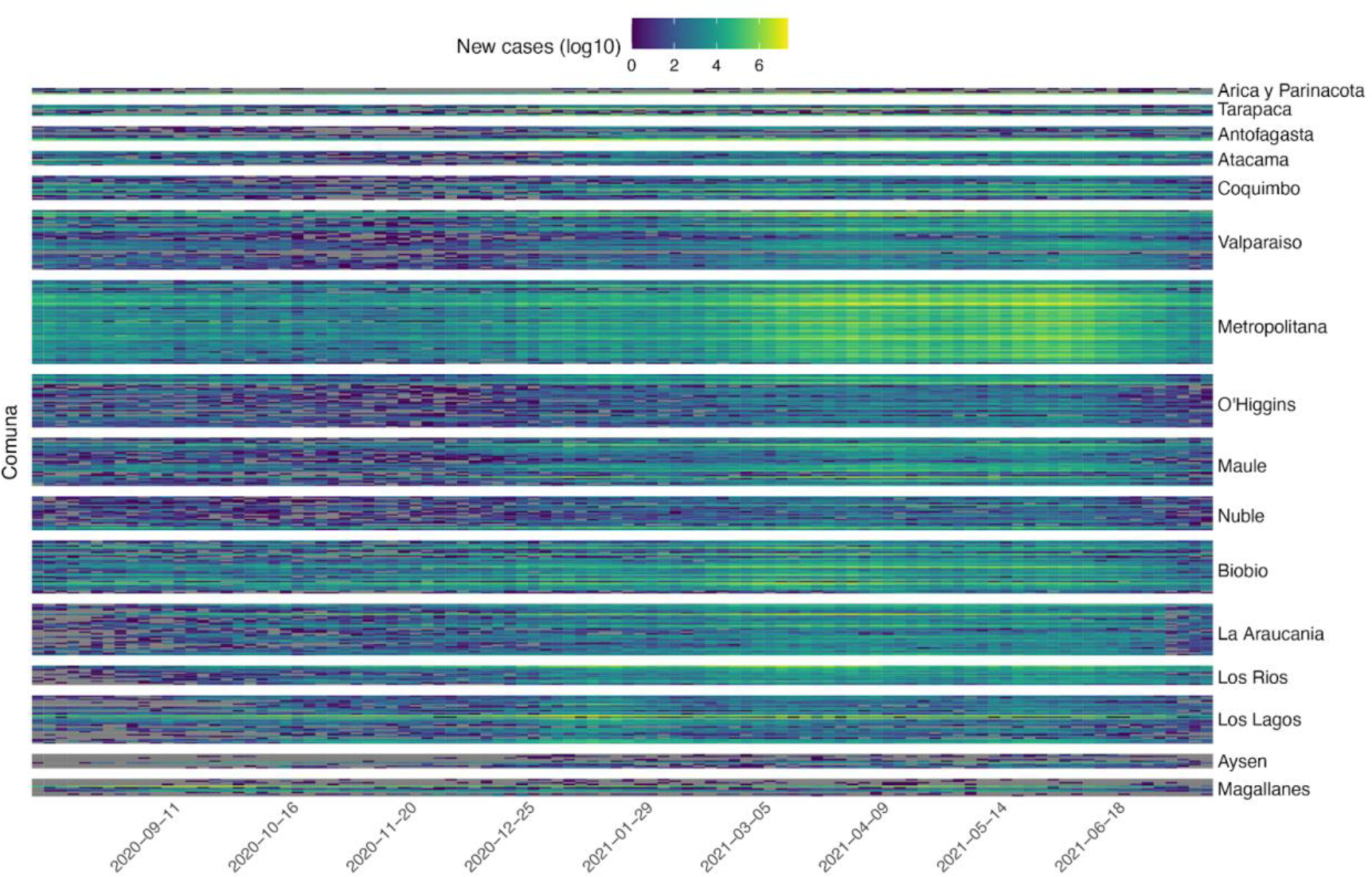
Reported new cases per comuna in Chile. New cases reported across Chile. Values correspond to dates when epidemiological reports were issued (every 2-4 days_ and do not represent a complete daily time series. Each row represents an individual comuna, and these are grouped by region (Adm1), ordered from north to south.

**Figure S8.**
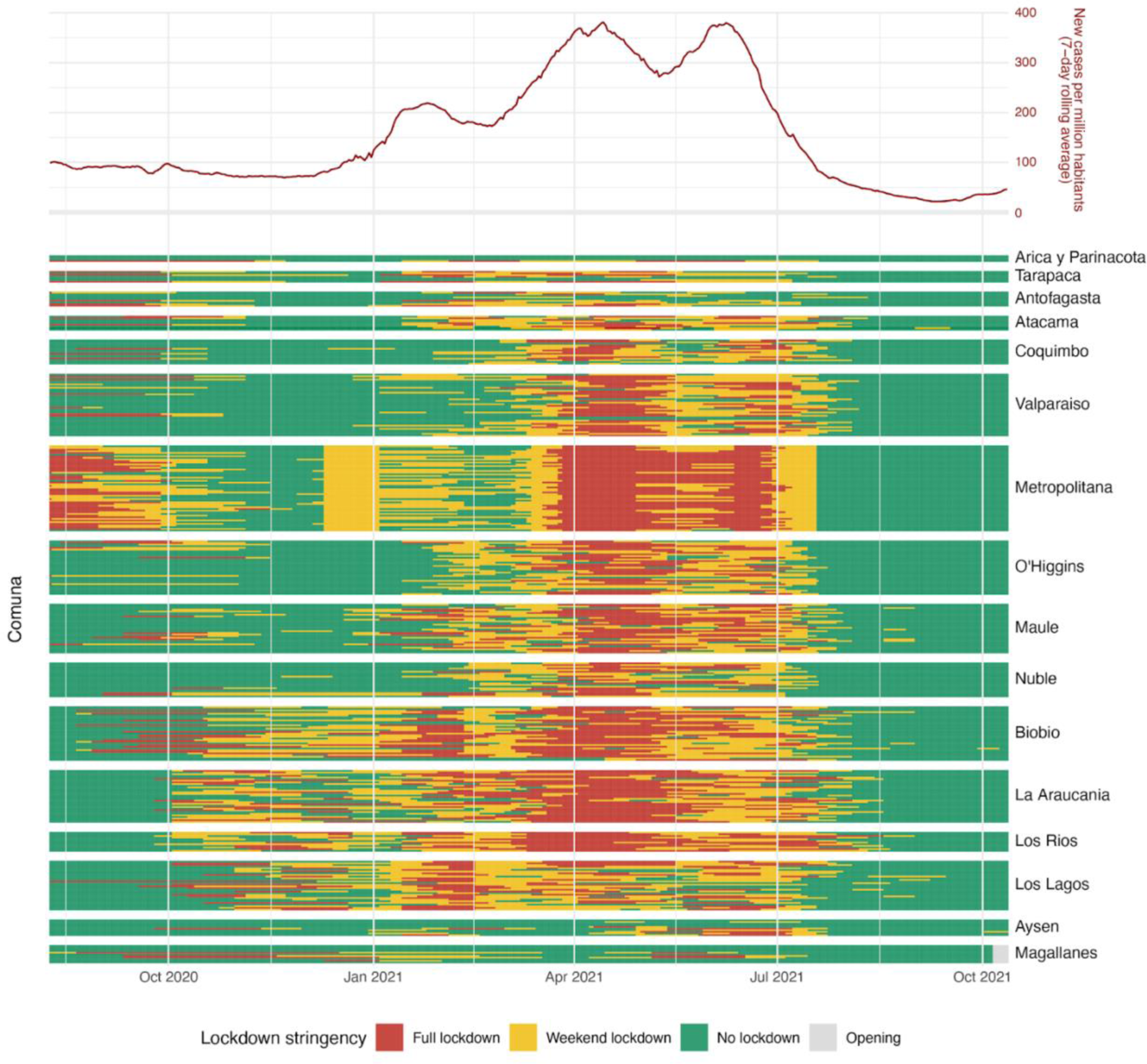
Changes of lockdown stringency in Chile. Upper panel shows the 7-day rolling average of total new cases in Chile (as reported by Our World in Data). The lower panel shows the daily lockdown stringency based on the number of weeks of the day where mobility restrictions were implemented within the Paso a Paso program. Details about lockdown stringency can be found in the Extended Text S1. Each row represents an individual comuna, and these are grouped by region (Adm1), ordered from north to south.

**Figure S9.**
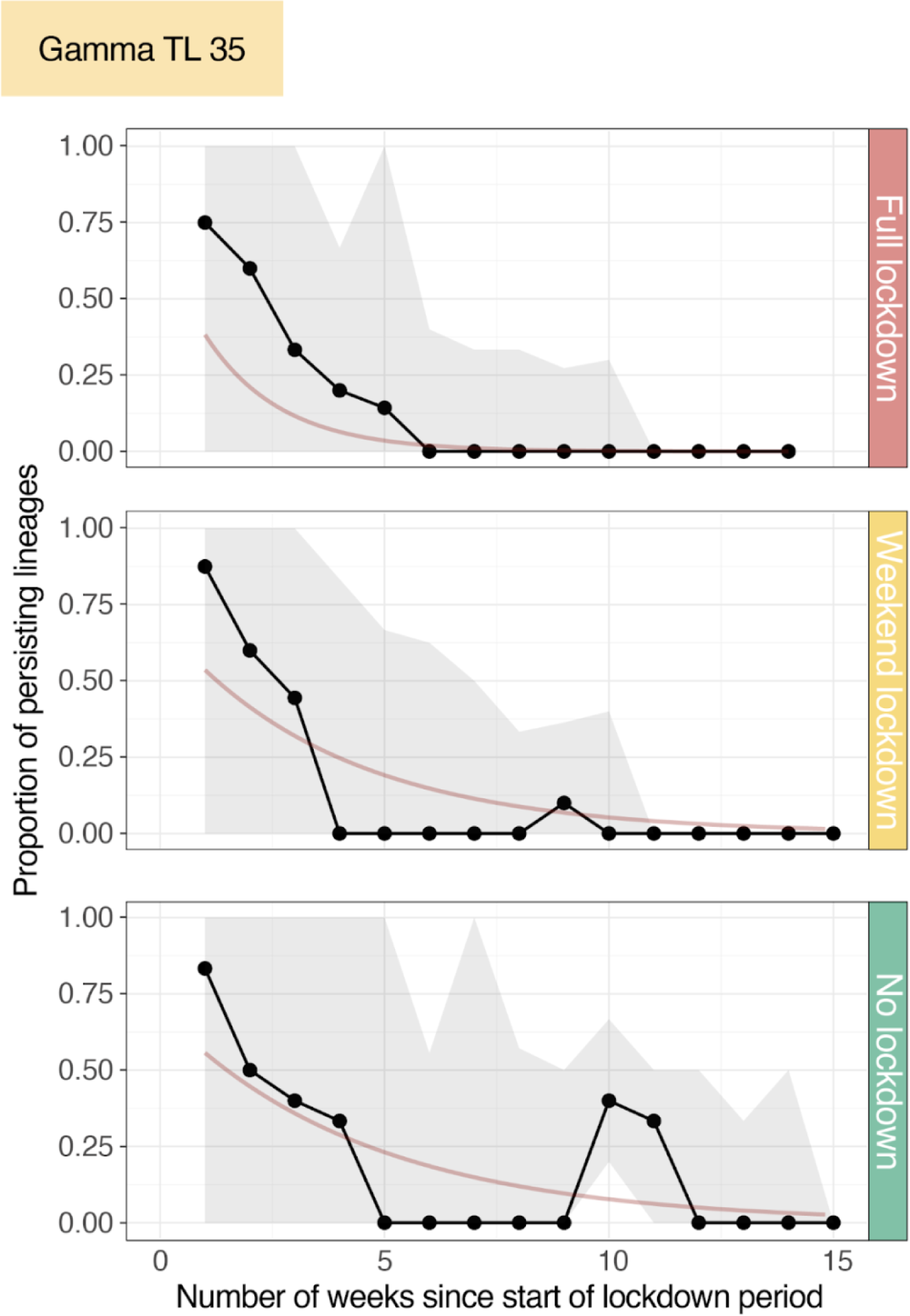
Decay of persisting viral lineages over time. Proportion of persisting lineages from Gamma TL 35 across the most affected comunas in Chile under different lockdown tiers estimated with PersistenceSummarizer. Persistence was measured on a weekly basis from the start of the lockdown period by calculating the proportion of phylogenetic branches (lineages) that persisted from the beginning of the lockdown. The dark red line shows a fitted exponential function to the median estimates of the persisting lineage proportions. Observations are aggregated over the fourteen comunas with the highest number of viral movements in our data set.

**Figure S10.**
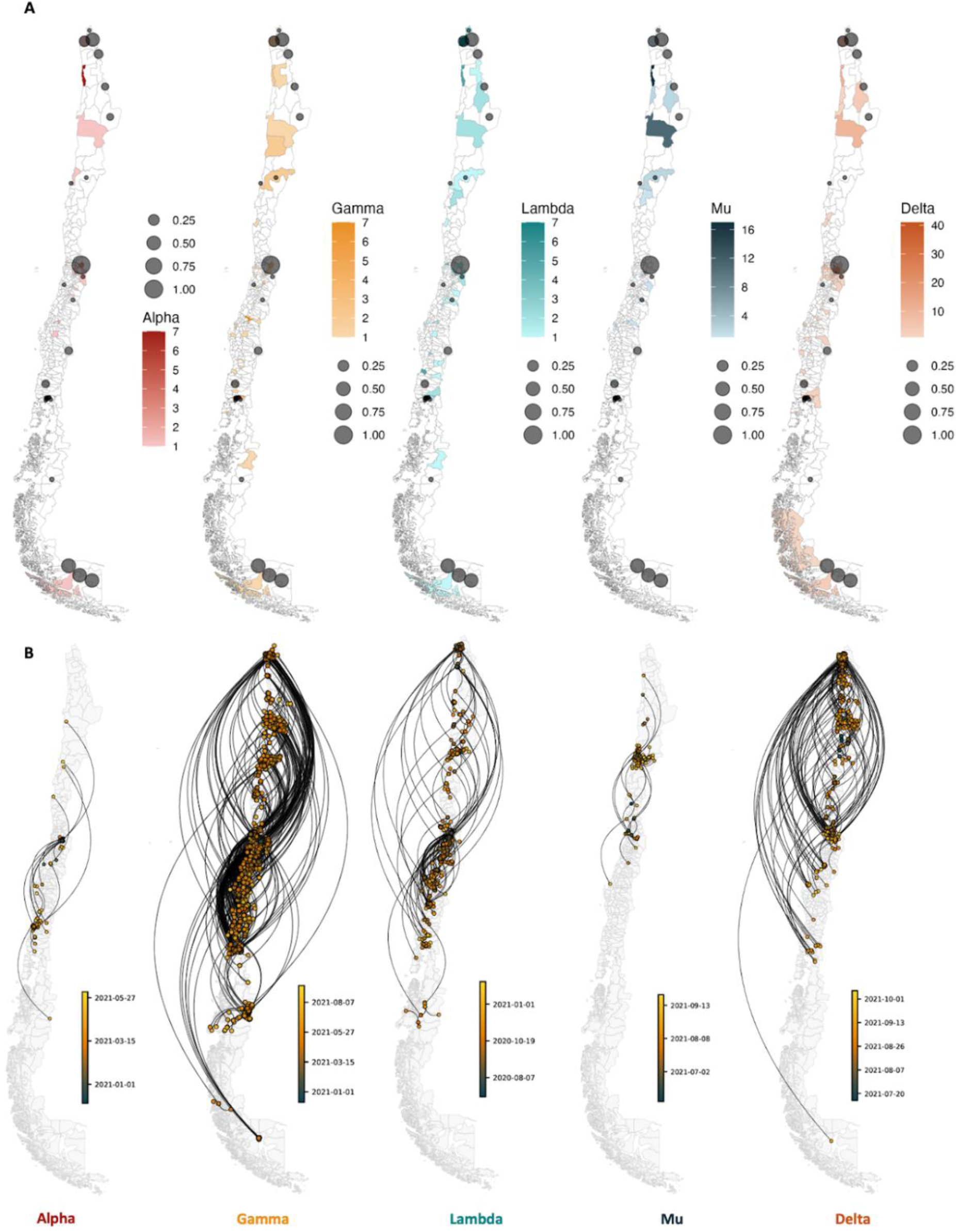
Number of first detection of SARS-CoV-2 introductions into Chile. Maps show the number of first detections of viral introductions (i.e., location of singletons and earliest sequences for each transmission lineage inferred from genomic data) by comuna (adm3) in Chile. Grey circles show the comunas where the main land border crossings in Chile are located, the radius shows the proportion of crossings that go through each one of these land border crossings.

**Figure S11.**
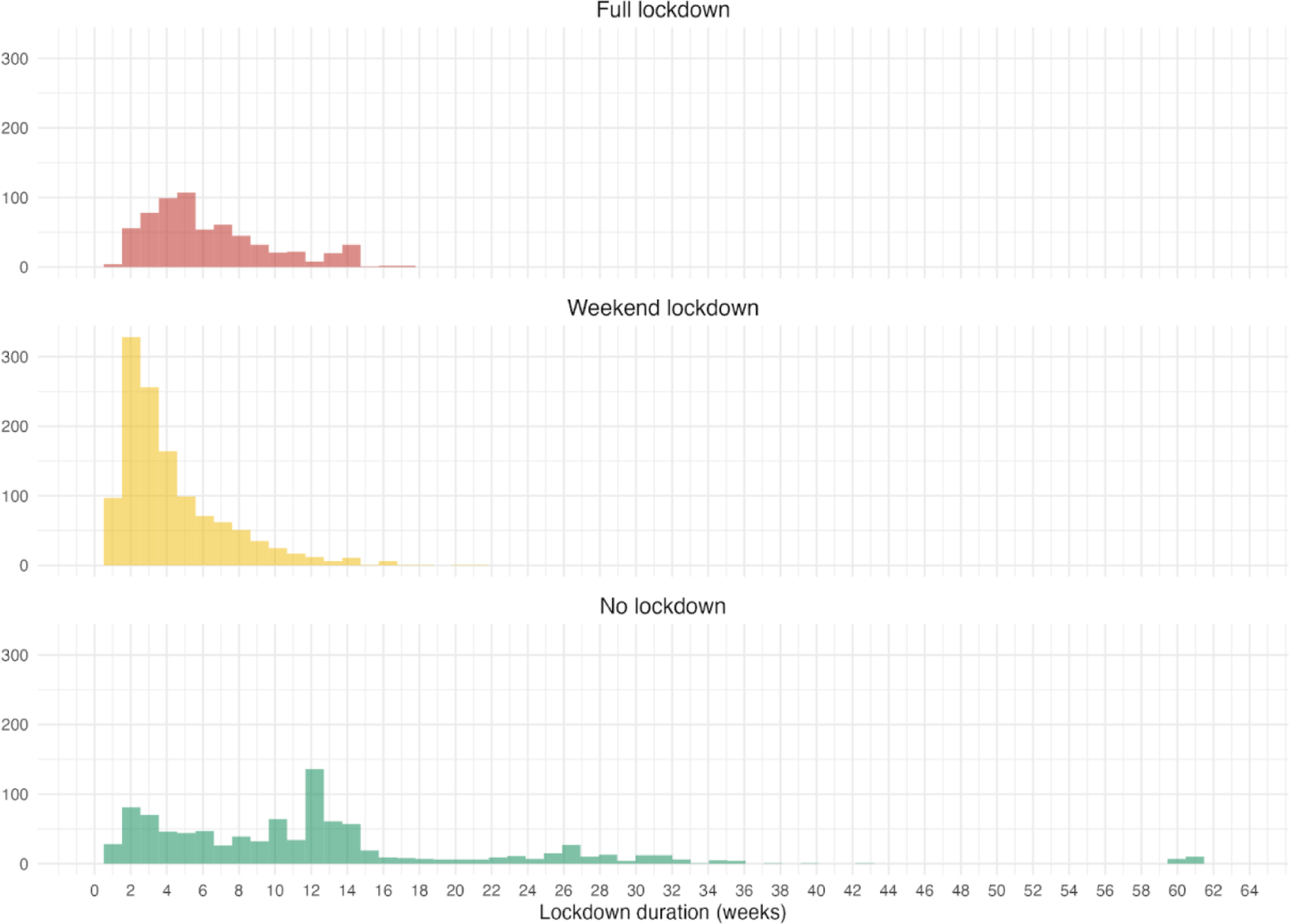
Duration of lockdown tiers in Chile. Distribution of the continuous number of weeks for which each lockdown tier was implemented. Every individual observation corresponds to a change of lockdown stringency tier in a single comuna, and the number of weeks are counted from the start of that stringency tier to the next change in the stringency tier for that comuna.

